# Systematic Review of Treatment of Beta-Cell Monogenic Diabetes

**DOI:** 10.1101/2023.05.12.23289807

**Authors:** Rochelle N. Naylor, Kashyap A. Patel, Jarno L.T. Kettunen, Jonna M.E. Männistö, Julie Støy, Jacques Beltrand, Michel Polak, ADA/EASD PMDI, Tina Vilsbøll, Siri A.W. Greeley, Andrew T. Hattersley, Tiinamaija Tuomi

**Affiliations:** Departments of Pediatrics and Medicine, University of Chicago, Chicago, Illinois, USA; University of Exeter Medical School, Department of Clinical and Biomedical Sciences, Exeter, Devon, UK; Helsinki University Hospital, Abdominal Centre/Endocrinology, Helsinki, Finland; Folkhalsan Research Center, Helsinki, Finland; Institute for Molecular Medicine Finland FIMM, University of Helsinki, Helsinki, Finland; Departments of Pediatrics and Clinical Genetics, Kuopio University Hospital, Kuopio, Finland; Department of Medicine, University of Eastern Finland, Kuopio, Finland; Steno diabetes center Aarhus, Aarhus university hospital, Aarhus, Denmark; APHP Centre Hôpital Necker Enfants Malades Université Paris Cité, Paris France; Inserm U1016 Institut Cochin Paris France; Department of pediatric endocrinology gynecology and diabetology, Hôpital Universitaire Necker Enfants Malades, IMAGINE institute, INSERM U1016, Paris, France; Université Paris Cité, Paris, France; American Diabetes Association/European Association for the Study of Diabetes Precision Medicine Initiative; Department of Clinical Medicine, University of Copenhagen; Lund University Diabetes Center, Malmo, Sweden

**Author notes:** These authors contributed equally (Kashyap A. Patel, Jarno L.T. Kettunen, Jonna M.E. Männistö, Julie Støy). These authors jointly supervised this work (Andrew T. Hattersley, Tiinamaija Tuomi). **Correspondence** Tiinamaija Tuomi.

**Keywords:** Precision Medicine, Monogenic Diabetes, Genetics, MODY, Neonatal Diabetes, Syndromic Diabetes, Mitochondrial diabetes, GCK, HNF1A, HNF4A, HNF1B, m.3243A>G, 6q24, SLC19A2

## Abstract

**Background:** Beta-cell monogenic forms of diabetes are the area of diabetes care with the strongest support for precision medicine. We reviewed treatment of hyperglycemia in GCK-related hyperglycemia, HNF1A- HNF4A- and HNF1B-diabetes, Mitochondrial diabetes (MD) due to m.3243A>G variant, 6q24-transient neonatal diabetes (TND) and SLC19A2-diabetes.

**Methods:** Systematic reviews with data from PubMed, MEDLINE and Embase were performed for the different subtypes. Individual and group level data was extracted for glycemic outcomes in individuals with genetically confirmed monogenic diabetes.

**Results:** 147 studies met inclusion criteria with only six experimental studies and the rest being single case reports or cohort studies. Most studies had moderate or serious risk of bias.

For GCK-related hyperglycemia, six studies (N=35) showed no deterioration in HbA1c on discontinuing glucose lowering therapy. A randomized trial (n=18 per group) showed that sulfonylureas (SU) were more effective in HNF1A-diabetes than in type 2 diabetes, and cohort and case studies supported SU effectiveness in lowering HbA1c. Two crossover trials (n=15 and n=16) suggested glinides and GLP-1 receptor agonists might be used in place of SU. Evidence for HNF4A-diabetes was limited. While some patients with HNF1B-diabetes (n=301) and MD (n=250) were treated with oral agents, most were on insulin. There was some support for the use of oral agents after relapse in 6q24-TND, and for thiamine improving glycemic control and reducing insulin requirement in SLC19A2-diabetes (less than half achieved insulin-independency).

**Conclusion:** There is limited evidence to guide the treatment in monogenic diabetes with most studies being non-randomized and small. The data supports: no treatment in GCK-related hyperglycemia; SU for HNF1A-diabetes. Further evidence is needed to examine the optimum treatment in monogenic subtypes.

## Introduction

In monogenic forms of diabetes, the underlying genetic cause has implications for the disease mechanism, treatment and prognosis. Defining the underlying genetic etiology also defines the pathophysiology resulting in hyperglycemia; this greatly increases the chances of finding an optimal specific therapy or therapies for glucose lowering. Pathogenic variations in single genes can either result in reduced insulin secretion as seen in the beta-cell subtypes or reduced insulin action in the insulin resistant subtypes. This systematic review relates to the treatment of beta-cell subtypes with the treatment of insulin resistant subtypes being reviewed elsewhere^1^.

Beta-cell monogenic forms of diabetes have been the area of diabetes care where there is the strongest support for a precision medicine approach for treating hyperglycemia^2,3^. The supporting evidence usually came from initial case reports leading to follow-up with case series and in some cases experimental studies/trials. The evidence base is considered strong in the commonest subtypes of monogenic diabetes such as glucokinase (GCK)-related hyperglycemia, Hepatic Nuclear Factor 1 alpha (HNF1A)-diabetes, and ATP sensitive potassium channel (KATP)-neonatal diabetes (ND, related to pathogenic variants in *KCNJ11* and *ABCC8*). Such evidence is yet to be shown for the more rare subtypes of monogenic diabetes. The varying levels of evidence combined with expert opinion has led to recommendation for optimum treatment in international guidelines such as International Society for Pediatric and Adolescent Diabetes (ISPAD)^4^, with the strongest support for sulfonylureas (SU) as first-line therapy for HNF1A-diabetes and Hepatic Nuclear Factor 4 alpha (HNF4A)-diabetes, no pharmacologic therapy for GCK-related hyperglycemia, insulin for Hepatic Nuclear Factor 1 beta (HNF1B)-diabetes and mitochondrial diabetes (MD) and high dose SU for KATP-ND. A key point is that these clinical guidelines were developed without systematic review of all the available evidence.

Systematic review will allow a comprehensive assessment of the strength of the evidence for the specific recommendations for precision medicine approaches. To our knowledge there is only one area of beta-cell monogenic diabetes where robust systematic reviews have been used-this is in the glycemic treatment of KATP-ND with high dose SU^5^ and the partial response of neurological features in KATP-ND to high dose SU therapy^6^. We therefore decided to systematically review the evidence of a precision medicine approach with an optimal glucose-lowering therapy for all the common subtypes of beta-cell monogenic diabetes except for the KATP-ND (Table 1). We restricted the analysis to the more common subtypes, as it is hard to have a sufficient number of individuals to determine optimum treatment in the less common forms.

**Table 1.** Subtypes of beta-call monogenic diabetes included in this systematic review.

| Monogenic diabetes subtype | Typical clinical features | Typical treatment approaches |
| --- | --- | --- |
| GCK-related hyperglycemia | Mild, stable hyperglycemia present from birth<br>Often incidentally diagnosed during routine clinical exams or during gestational diabetes screening in pregnancy<br>Fasting blood glucose typically ranges between 5.5-8.0 mmol/L and HbA1c between 5.6-7.8% | No pharmacologic treatment |
| HNF1A-diabetes | Progressive insulin secretory defect with onset of diabetes typically in the second and third decades of life<br>Reported to have a lowered renal threshold for glycosuria, which may be an early sign | Sensitive to sulfonylureas |
| HNF4A-diabetes | Progressive insulin secretory defect with onset typically in the second and third decades of life<br>May have fetal macrosomia and hyperinsulinemic hypoglycemia in the neonatal and early-childhood period | Sensitive to sulfonylureas |
| HNF1B-diabetes | Syndromic form of diabetes with onset in the second and third decades of life, but typically later than in HNF1A-diabetes and HNF4A-diabetes<br>Often with renal cysts or other developmental renal disease (single kidney, horseshoe kidney)<br>Other features can include hypoplasia of the pancreas, pancreatic exocrine deficiency, genital tract and biliary abnormalities, hypomagnesemia and neurodevelopmental disorders (with whole-gene deletions) | Use of OHA reported, most cases insulin-treated |
| m.3243A Mitochondrial Diabetes | Maternally-inherited syndromic form of diabetes, often with sensorineural deafness<br>Diabetes typically occurs in the 30s but onset ranges from 11-68 years<br>Other features can include cardiomyopathy, myopathy, epilepsy, lactic acidemia, macular dystrophy, renal disease (e.g., focal segmental glomerular sclerosis) | Use of OHA reported, most cases insulin-treated |
| 6q24 Transient Neonatal Diabetes | Neonatal onset of diabetes usually within the first week of life, typically associated with severe intrauterine growth restriction and small for gestation age at birth<br>May have macroglossia and umbilical hernia<br>Diabetes resolves by 18 months of age (average duration is 3 months)<br>Diabetes may relapse in adolescence, or pregnancy (times of increased insulin resistance) or later in adulthood | Insulin or SU in neonatal phase<br>Various glucose-lowering treatments in relapse phase |
| SLC19A2-diabetes | Diabetes onset is often in the infancy period, but can occur later in childhood or adolescence<br>Megaloblastic anemia is responsive to treatment with thiamine<br>Other main features include sensorineural deafness | Thiamine for anemia, most cases insulin-treated |

This systematic review is written on behalf of the American Diabetes Association (ADA)/European Association for the Study of Diabetes (EASD) *Precision Medicine in Diabetes Initiative* (PMDI) as part of a comprehensive evidence evaluation in support of the 2^nd^ International Consensus Report on Precision Diabetes Medicine^7^. The PMDI was established in 2018 by the American Diabetes Association (ADA) in partnership with the European Association for the Study of Diabetes (EASD) to address the burgeoning need for better diabetes prevention and care through precision medicine^8^. The evidence for whom to test for monogenic diabetes, how to test them and how to interpret a gene variant, as well as for underpinning the link between the genetic test result and prognostics are covered as separate systematic reviews in this series, by other members of the PMDI addressing precision diagnostics and prognostics for monogenic diabetes^9,10^.

### Aims

The specific areas where we aimed to provide a systematic review of the evidence for precision medicine in beta-cell monogenic diabetes were:

1. What is the optimal glucose lowering therapy in the three commonest subtypes of autosomal dominant familial diabetes also known as Maturity Onset Diabetes of the Young (MODY): GCK-related hyperglycemia, HNF1A-diabetes and HNF4A-diabetes?
2. What is the optimal glucose lowering therapy in the two commonest subtypes of syndromic diabetes: HNF1B-diabetes and Mitochondrial Diabetes (MD) due to m.3243A>G in the MT-TL1 gene?
3. Are there alternatives to insulin therapy in 6q24 transient neonatal diabetes (6q24-TND); and in SLC19A2-diabetes also known as Thiamine-Responsive Megaloblastic Anemia (TRMA) does thiamine supplementation improve glycemia in TRMA syndrome?

## Methods

Protocols for systematic reviews were developed and registered in Prospero (https://www.crd.york.ac.uk/prospero/; for MODY (CRD42021279872); syndromic diabetes (CRD42021250955) and neonatal diabetes (CRD42023399408). The study was reported in accordance with Preferred Reporting Items for Systematic Reviews and Meta-Analysis (PRISMA) guidelines (http://prisma-statement.org).

### Search strategy

We comprehensively reviewed the literature associated with glycemic treatment outcomes, searching in PubMed, MEDLINE and Embase separately for 1) GCK-related hyperglycemia, HNF1A-diabetes and HNF4A-diabetes; 2a) HNF1B-diabetes; 2b) MD with m.3243A>G variant; 3a) 6q24-TND; 3b) SLC19A2-diabetes. The OMIM (and ORPHA) codes for the diseases are 1) #125851 (552), #600496 (552), #125850 (552), respectively; 2a) #137920 (552); 2b) #520000 (225); 3a) #601410 (99886), and 3b) #249270 (49827).

Filtering and selection of studies for full text review and data extraction were recorded using Covidence (https://www.covidence.org). The search strategies are shown in the supplemental tables and the PRISMA summaries for the searches are shown in Figures 1-4. At least two authors independently reviewed the titles and abstracts to filter out the articles for full-text review. Two authors independently reviewed full text articles and either a third author (regarding GCK-related hyperglycemia, HNF1A-diabetes and HNF4A-diabetes) or the two reviewers jointly (HNF1B-diabetes, MD, 6q24-TND, SLC19A2-diabetes) resolved discrepancies. One author for each search performed the data extraction using a standardized form and two reviewed it. Data were extracted from text, tables or figures from the main and supplementary documents. Data extracted for each study included first author, publication year, country, study design, number of participants, age at diagnosis of diabetes and at the time of study, sex, diabetes duration, treatment information, and glycemic data. Figures 1-4 show the flow-charts for the searches.

**Figure 1.**
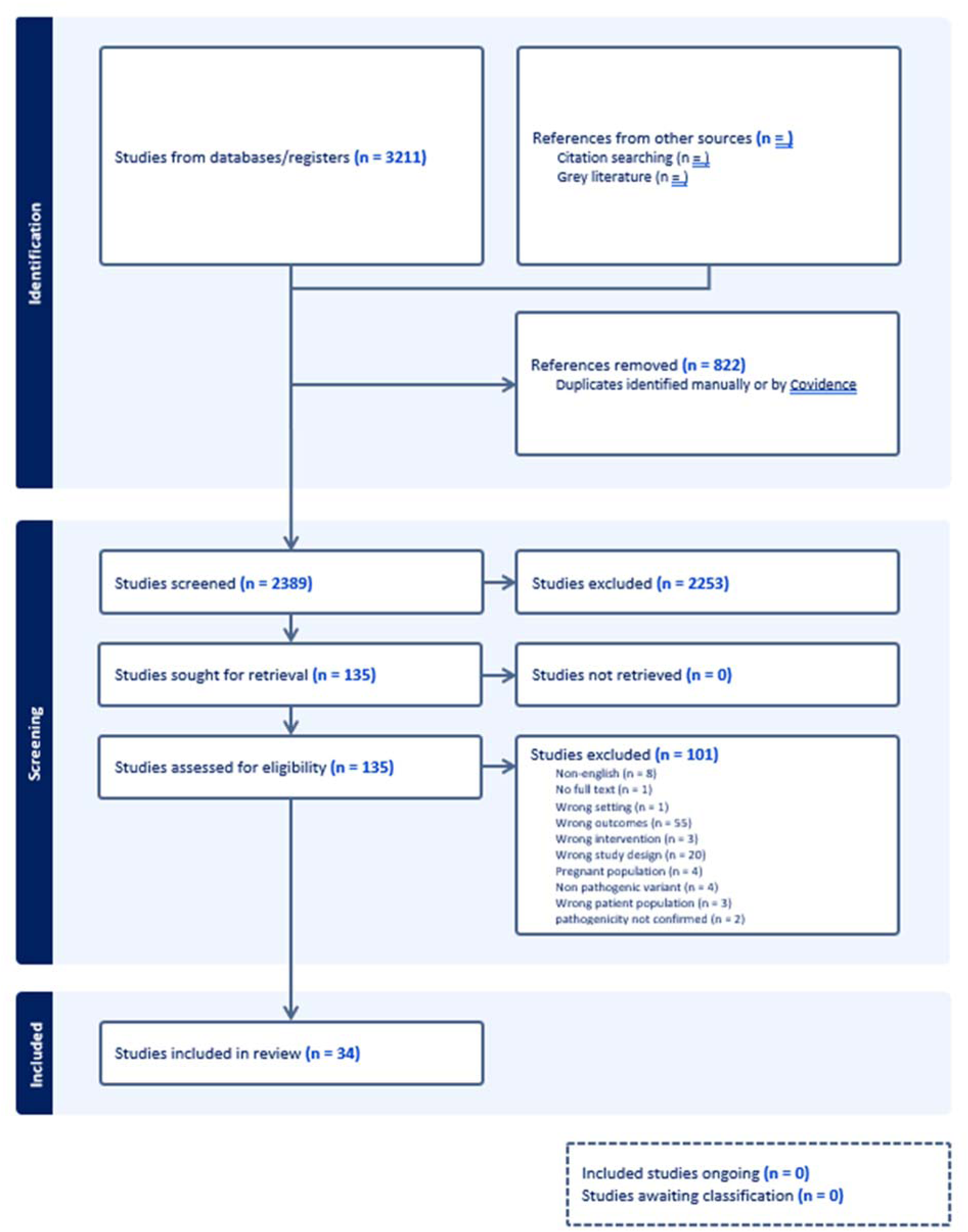
PRISMA search summary for GO<-, HNFlA- and HNF4A MOD.

### Inclusion and exclusion criteria

We included English language original articles (case reports, case series, cross-sectional studies, experimental studies, trials) written after 1994 (following the initial molecular characterization of monogenic causes of diabetes) concerning treatment of hyperglycemia in human individuals diagnosed with the monogenic diabetes subtypes of interest. Individuals with variants of unknown significance, multiple types of diabetes, and those lacking measures of treatment effect were excluded. Studies or data within studies that aggregated treatment effects for multiple monogenic diabetes types were also excluded. We included studies reporting on more than one monogenic diabetes type of interest with partially incomplete data, as long as data could be fully extracted for at least one monogenic diabetes subtype.

### Risk of bias and evidence appraisal

We used NHLBI study quality assessment tools to assess risk of bias (https://www.nhlbi.nih.gov/health-topics/study-quality-assessment-tools) and the methods outline by Sherifali et al, to assess the level of evidence and to grade recommendations^11^.

### Data synthesis

Data were extracted from Covidence into Microsoft Excel. Data were summarized using Microsoft Excel. Data are presented as mean ± standard deviation, median [interquartile range, IQR], median (range) or as percentages.

Meta-analysis was not carried out due to the heterogeneity in study design as well as the high level of risk of bias in included studies, particularly case reports and series that make up a substantial portion of the literature.

## Results

### Risk of bias

Most studies across all diabetes types were rated as having moderate or serious risk of bias related to the study design (mainly comprising observational case reports and case series) and selection of the study population and outcome variables (with genetic diagnoses not based on non-targeted population screening). Whenever a response or a non-response to a drug was reported, it was unclear whether other factors significantly or moderately contributed to the reported response.

#### 1. What is the optimal glucose lowering therapy in the three commonest subtypes of autosomal dominant familial diabetes (MODY): GCK-related hyperglycemia, HNF1A-diabetes and HNF4A-diabetes?

There were 2389 studies identified by the literature search (Figure 1). After duplicate removal and title and abstract review, 136 studies remained for full text review. Data was extracted from 34 articles in total, including six experimental studies, of which four were randomized controlled studies, 26 case reports, series or cohort studies, and two cross-sectional studies, one of which also presented cohort data. There were 13 studies that contributed to data on treatment for GCK-related hyperglycemia, 22 studies for HNF1A-diabetes, and three studies for HNF4A-diabetes. The key summaries of these data are in Tables 2 and 3. The overall level of evidence was low and the risk of bias high for most studies.

**Table 2.** Experimental studies of GCK-related hyperglycemia and HNF1A-diabetes.

| Study Name, Design & Population | Primary Endpoint | Key Results(s) | Appraisal of Study |
| --- | --- | --- | --- |
| **GCK-related hyperglycemia** |  |  |  |
| **Klupa 2011**<br>Observational crossover study of 2 days of high, followed by 2 days of low carbohydrate intake with a 1-day washout period<br>10 adults (4 females) with GCK-related hyperglycemia<br>Mean age 37.4 (range 19-54) years (diabetes, N=7), 30.0 (19-52) years (prediabetes, N=3)<br>Mean baseline HbA1c 7.3 (6.1-8.4)% (diabetes), 6.0 (5.9-6.1)% (prediabetes) | -Mean blood glucose level (MBG)<br>-Percentage of time above target postprandial blood glucose level defined as 7.8 mmol/l | When the 3 patients with prediabetes were excluded, the remaining 7 patients had higher MBG during exposure to the high-carbohydrate diet than while on the low-carbohydrate diet (difference of 0.76 mmol/l, P = 0.02)<br>They also had spent less time above the target postprandial blood glucose level (difference of 11.7%, P = 0.02) | -Lacked a control or comparison group<br>-Did not control for carryover effects<br>• Short washout period<br>-Did not control for potential order effects<br>• All participants began with high-carbohydrate diet<br>-Diet and daily caloric intake was not standardized<br>-In analysis, groups were divided into diabetes and prediabetes based on FBG. This resulted in overlap of HbA1c between groups and individuals with HbA1c values that are defined as prediabetes analyzed in the diabetes group.<br>**With study design limitations, short duration, and separate analysis of prediabetes from diabetes, which was not a predefined exclusion, no recommendation can be made on dietary treatment for GCK-related hyperglycemia.** |
| **HNF1A-MODY** |  |  |  |
| **Pearson 2003**<br>Randomized, crossover trial beginning with a 1-week washout period off treatment before 6 weeks of either gliclazide or metformin followed by 6 weeks of metformin or gliclazide with a 1-week washout period<br>18 adults (11 females) with HNF1A-diabetes*<br>Mean (SD) age 44 years (13) and diabetes duration 18.3 years (9.2)<br>Mean (SD) FPG 12.1 mmol/L (2.6)<br>Mean (SD) Fructosamine 450 umol/L (104) | -Reduction in fasting plasma glucose (FPG) | -Participants with HNF-1A diabetes had a 5.2-fold greater response to gliclazide than to metformin (FPG reduction of 4.7 versus 0.9 mmol/L, respectively).<br>-Participants with type 2 diabetes had no difference in the response to gliclazide and metformin (FPG reduction of 1.2 and 1.3 mmol/L, respectively).<br>-The fall in FPG in response to gliclazide was 3.9-fold greater in HNF1A-diabetes compared to type 2 diabetes.<br>-There was no statistically significant difference in the fall in FPG in response to metformin between HNF1A-diabetes and type 2 diabetes. | -Small sample size and short duration are limitations of this study<br>-Strengths include study type (controlled experiment), intent-to-treat analysis and assessing for carryover effects<br>**This study provides evidence to establish SU as a precision treatment for HNF1A-diabetes, showing treatment response differences by genotype.** |
| 18 adults (6 females) with type 2 diabetes*<br>Mean (SD) age 66 years (6) and diabetes duration 4.8 years (3.6)<br>Mean (SD) FPG 10.5 mmol/L (3.1)<br>Mean (SD) Fructosamine 378 μmol/L (105)<br>*Matched by body-mass index and FPG after the washout period off treatment |  | -In HNF1A diabetes, glycemia, measured by fructosamine, was 136 μmol/L lower on gliclazide than on metformin.<br>-In type 2 diabetes the fructosamine was 23 μmol/L higher on gliclazide than metformin. |  |
| **Tuomi 2006**<br>Randomized, double-blinded, crossover study beginning with a washout period off treatment (differed by baseline diabetes regimen) and then comparing the acute effect of nateglinide, glibenclamide, and placebo in conjunction with a standard 450-kcal test meal and light bicycle exercise with a minimum 1-week washout between treatments<br>15 adults (10 females) with HNF1A-diabetes<br>Median (IQR) age 41 years (16.5)<br>Median HbA1c 6.7% (2.3) | -Prandial plasma glucose concentrations<br>• Peak plasma glucose<br>• Maximum increment in plasma glucose from fasting to 140 minutes<br>• Incremental glucose area under the curve (AUC) in the first 140 minutes of the test | -Peak plasma glucose and incremental glucose AUC remained significantly lower at the nateglinide visit compared with glibenclamide or placebo.<br>-During exercise, there were 6 episodes of hypoglycemia in the glibenclamide group compared to none with nateglinide and placebo (P = 0.030 for both comparisons). Three patients had to interrupt the test due to hypoglycemia. | -This is not a true treatment study as individuals only received a single dose of each medication<br>-There were different washout periods at baseline and during the study<br>**Although not a true treatment study, this is the only controlled study providing evidence for the use of glinides as an alternative to SUs when hypoglycemia is a concern and supports the limited case reports citing beneficial use of glinides in these circumstances.** |
| **Østoft 2014**<br>Randomized, double-blinded, placebo-controlled, crossover study beginning with 1 week washout period off treatment followed by 6 weeks of liraglutide and placebo, followed by 6 weeks of glimepiride and placebo with a 1-week washout<br>Or<br>6 weeks of glimepiride and placebo followed by 6 weeks of liraglutide and placebo with a 1-week washout<br>16 adults (8 females) with HNF1A-diabetes<br>Mean (range) age 39 years (23-67)<br>Mean (SD) HbA1c 6.4% +/- 0.2 | -FPG measured at baseline and at the end of each treatment | -Both treatments resulted in significantly lower FPG compared with baseline but did not statistically differ from each other (P = 0.624).<br>**Secondary endpoint**<br>-A total of 19 mild hypoglycemic events were reported in 11 participants. One event was reported during treatment with liraglutide and 18 events during treatment with glimepiride. | -Small sample size and short duration are limitations of this study<br>-Treat to target (FPG 5.0-5.9 mmol/L) design may have contributed to hypoglycemic events with glimepiride treatment and so may overcall differences that would be seen in a non-study setting<br>**This study provides evidence of short-term efficacy of GLP1RA (liraglutide) in treatment of HNF1A-diabetes. Larger studies with a longer duration allowing comparison of the HbA1c response are needed, particularly given a trend of a larger FPG decrement from baseline with glimepiride (-2.8 +/- 0.7 mmol/L) than liraglutide (-1.6 +/- 0.5 mmol/L)** |
| **Christensen 2020**<br>Randomized, double-blinded, placebo-controlled, crossover study of 16 weeks of glimepiride + linagliptin, followed by 16 weeks of glimepiride + placebo with a 4-week washout period<br>Or<br>16 weeks of glimepiride + placebo, followed by 16 weeks of glimepiride + linagliptin with a 4-week washout period<br>19 adults (11 females) with HNF1A-diabetes<br>Mean (SD) age 43 (14) years<br>Median diabetes duration 20 (range 8-34) years<br>Mean HbA1c 7.4% (0.7) | -Mean amplitude of glycemic excursions (MAGE), measured via 6 days of continuous glucose monitoring at the end of each treatment period | -Reduction in MAGE did not differ significantly between the two groups.<br>**<u>Secondary endpoints</u>**<br>-A -0.5% [-0.9 to -0.2] reduction in HbA1c was observed with glimepiride + linagliptin compared with glimepiride + placebo (P = 0.0048).<br>-15 vs. 32 episodes of hypoglycemia on CGM were observed with glimepiride + linagliptin versus glimepiride + placebo. | -Small sample size and relatively short duration are limitations of this study<br>-Strengths include intent-to-treat analysis and assessing for carryover effects and the impact of subject relatedness on results<br>**While larger studies of longer duration are needed, this study supports DPP4i (linagliptin) as add-on therapy to SU. Frequently, SU dose could be lowered with adding linagliptin and this was a weight neutral approach compared to 1.2 kg weight gain with SU monotherapy.** |

##### a) GCK-related hyperglycemia

We originally posed the question of what is the impact of pharmacological and non-pharmacological glucose lowering therapy in GCK-related hyperglycemia. However, we identified only one case report that presented data on active pharmacological intervention^12^ and one study on dietary intervention^13^ and the rest of the 10 studies^14–23^ assessed the impact of stopping anti-hyperglycemic agents on HbA1c or glucose or assessed the stability of HbA1c over time on no therapy (Table 3). Thus, we concentrated on analyzing the evidence to support or refute non-treatment of GCK-related hyperglycemia.

**Table 3.**
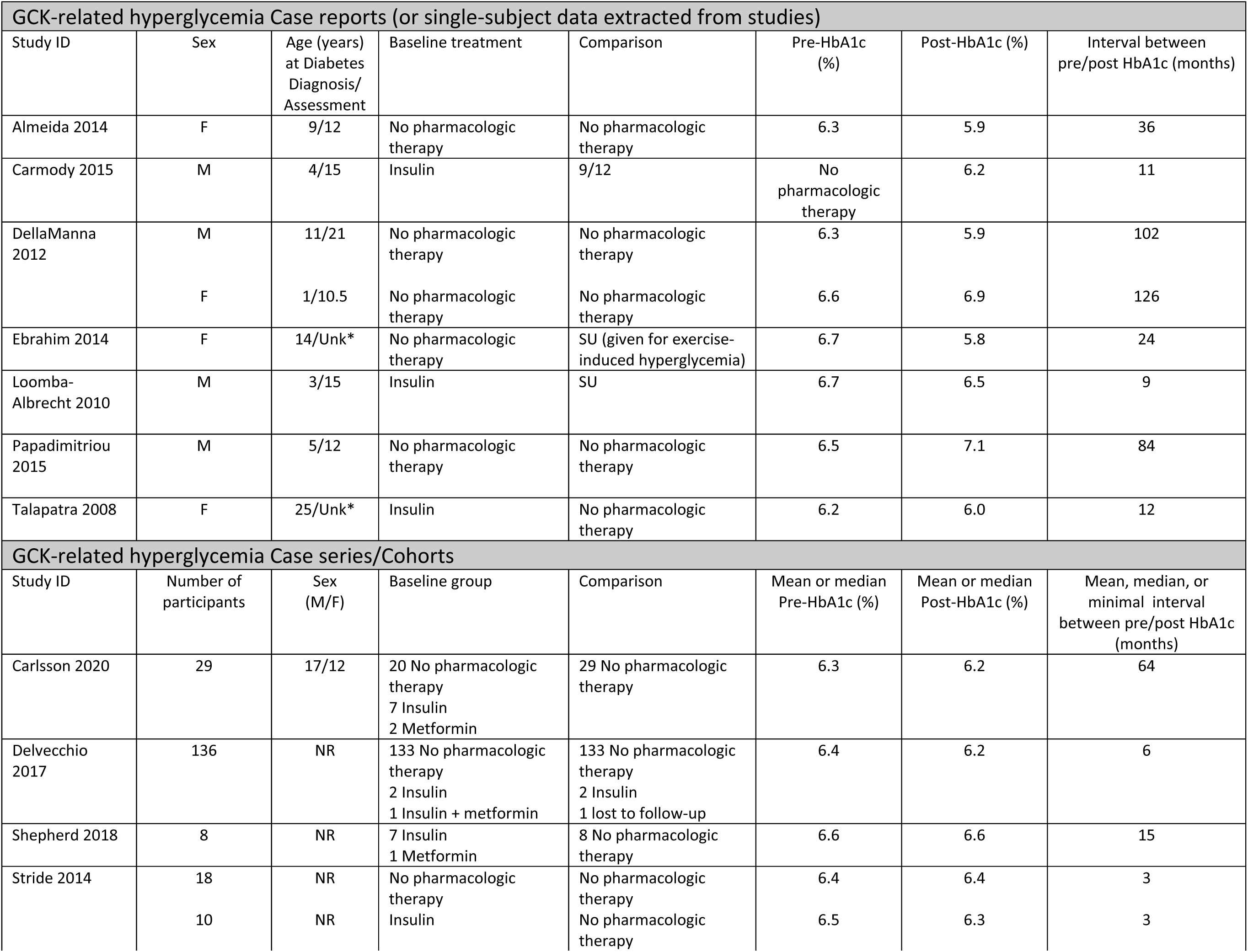
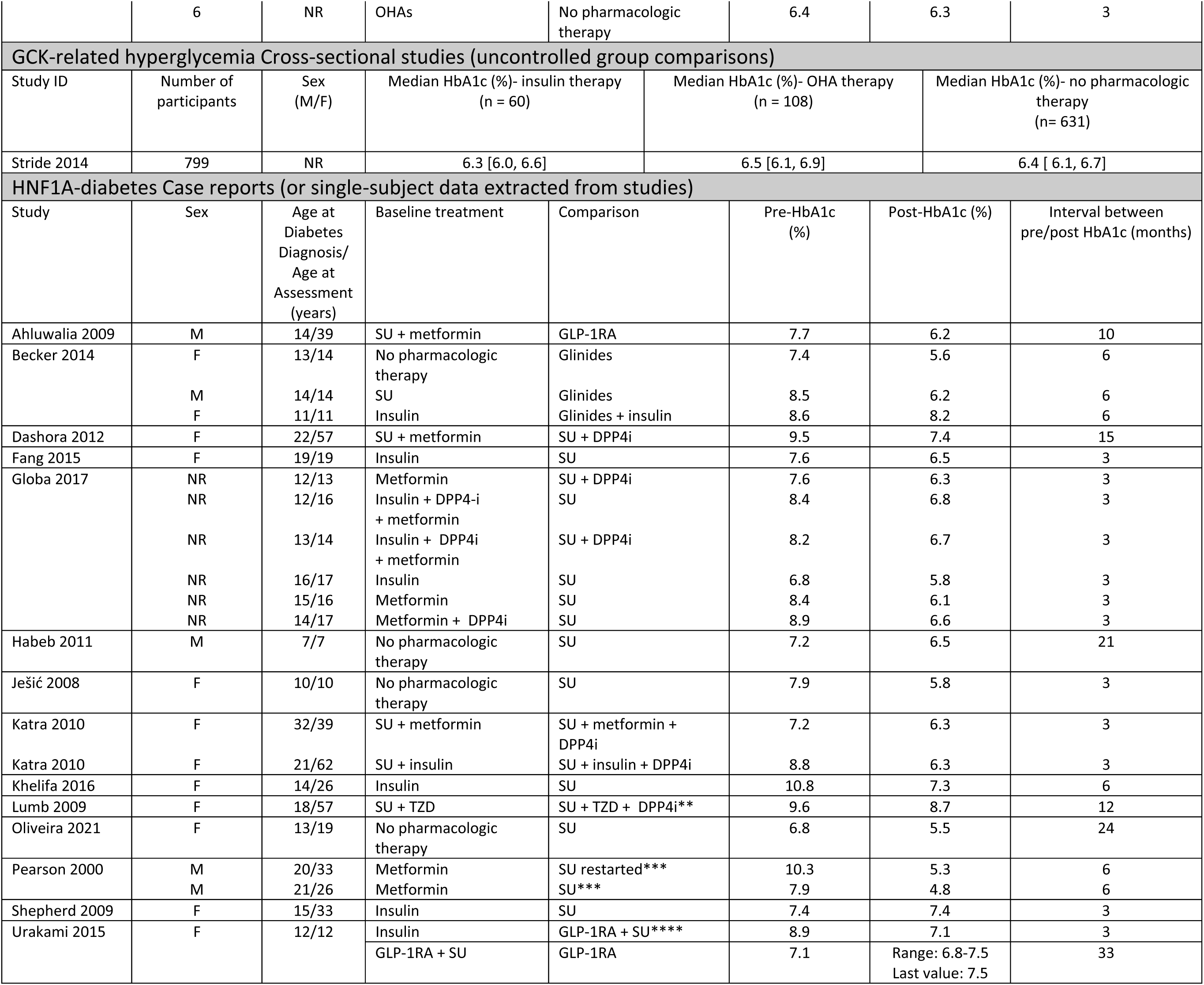
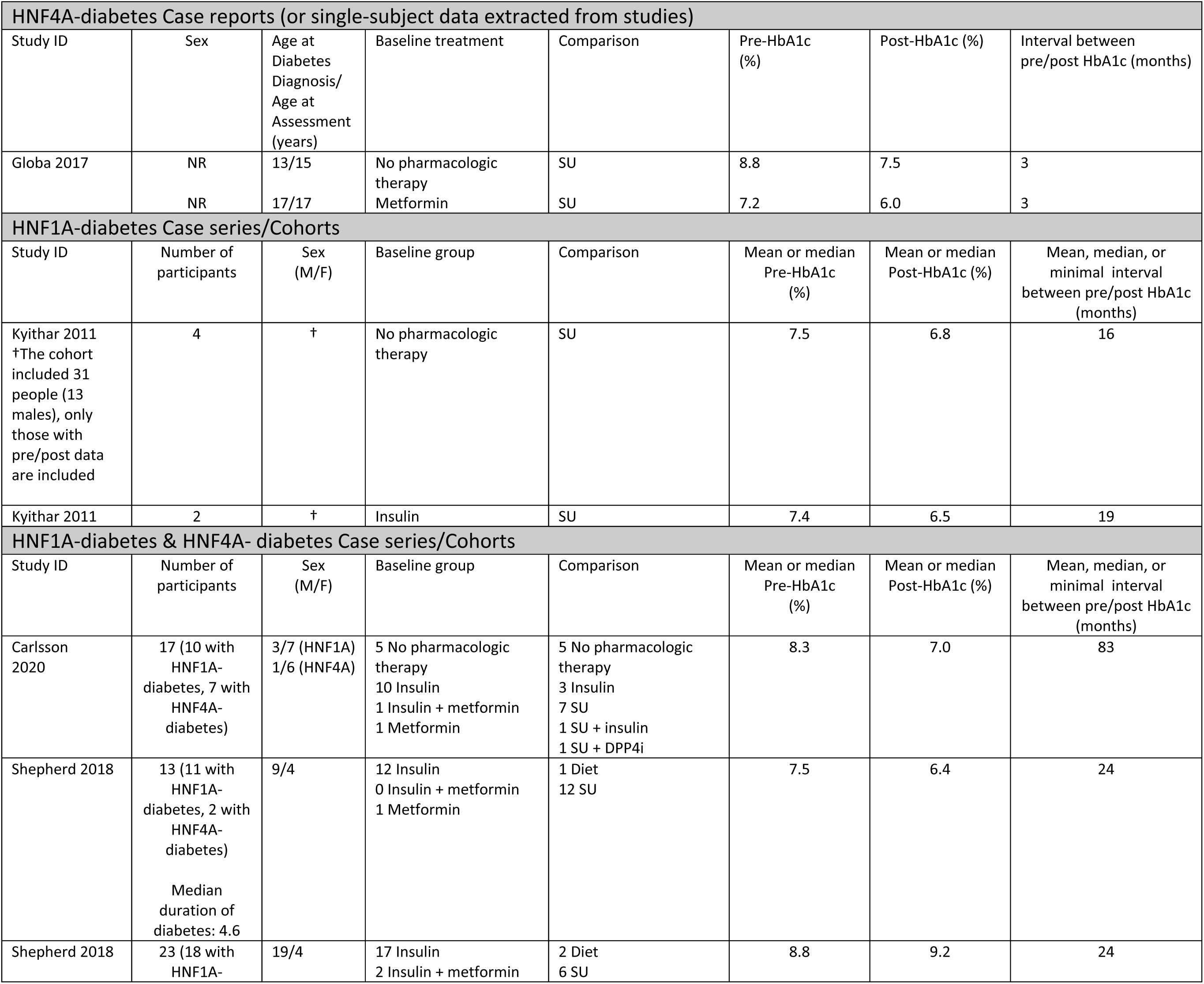
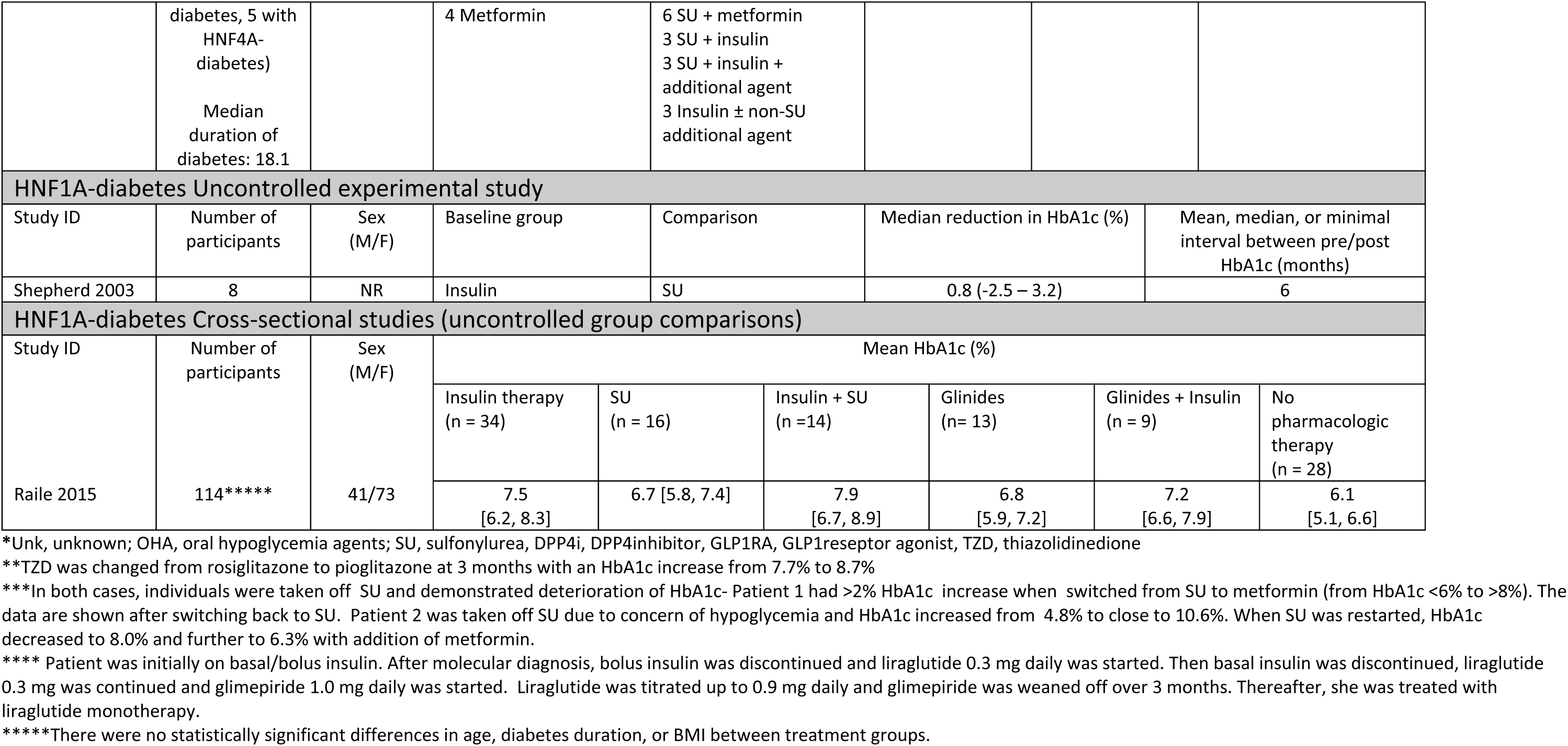
Summary of included studies for GCK-related hyperglycemia, HNF1A-diabetes, and HNF4A-diabetes.

Most studies on GCK-related hyperglycemia looked at within individual stability of HbA1c over time without glucose lowering therapy (175 individuals from case reports and cohort data, range of time between 3-126 months)^14,16,18,20–21,23^ or after cessation of pharmacologic therapy following genetic diagnosis (35 individuals from case reports and cohort data)^15,19–20,22–23^. All studies showed stability of HbA1c with no significant change including when glucose lowering therapy of either oral hypoglycemic agents (OHA) or insulin was discontinued (table 3). Only a single case report suggested adding therapy might lower HbA1c^12^.

A single large cross-sectional cohort study (n=799) showed no differences in HbA1c between observational non-randomized treated and untreated groups^23^.

There were no randomized long term treatment trials in GCK-related hyperglycemia. We identified a single experimental crossover study for GCK carriers^13^ that assessed the impact of high-carbohydrate (60%) versus low-carbohydrate (25%) unstandardized diet on mean glucose levels (MBG) and time spent above target postprandial blood glucose level (7.8 mmol/L) as measured by continuous glucose monitor (CGM) in 10 GCK subjects (Table 2). The duration of each intervention was brief (2 days with 1 day washout period). A statistically significant difference in mean glucose level (0.78 mmol/L) and time above target (11.7%) was found after high-carbohydrate diet if the analysis was restricted to the seven patients with an initial HbA1c above 6.5% (a non-predefined analysis). No comparison groups such as individuals without diabetes or with type 2 diabetes were included.

##### b) HNF1A-diabetes and HNF4A-diabetes

We analyzed studies for evidence of SU as an effective glucose-lowering therapy for HNF1A-diabetes and HNF4A-diabetes. We also assessed evidence for alternative or augmentative non-insulin therapies.

A striking observation was that all the trials or experimental studies (4 trials, 1 uncontrolled study, 76 individuals)^24–28^ and almost all the observational data (18 studies, 182 individuals)^20,22,29–44^ was for HNF1A-diabetes. While five studies included both HNF1A-diabetes and HNF4A-diabetes, only three studies (16 individuals) had HNF4A-diabetes data that could be extracted^20,22,33^.

In HNF1A-diabetes a single randomized cross-over study tested using SU in comparison with type 2 diabetes^24^ (Table 2). This was the only study in monogenic beta-cell diabetes to have a comparative group with type 2 diabetes, hence truly testing if it was a specific precision approach. The study compared the fasting plasma glucose (FPG) response in individuals with HNF1A-diabetes (N=18) and type 2 diabetes (N=18) to therapy with SU (gliclazide) or metformin for 6 weeks. The study groups were matched for body mass index and FPG after the wash-out period off-treatment but not for sex or age. Gliclazide was superior to metformin in lowering fasting glucose in HNF1A-diabetes but not in type 2 diabetes (5-fold greater response to SU than metformin in HNF1A-diabetes; no difference in type 2 diabetes).

Glinides have been proposed as an alternative to long-acting SU, for example in instances of problematic hypoglycemia. There was one randomized, placebo-controlled cross-over study that compared the acute effects of premeal dosing of SU (1.25 mg glibenclamide), glinide (30 mg nateglinide) or placebo on glucose excursions during and after a standardized meal and exercise in 15 participants^25^ (Table 2). When comparing glibenclamide and nateglinide response to the standardized meal, peak insulin occurred earlier and plasma glucose levels (peak and up to 140 min) were lower with nateglinide. Exercise after the meal resulted in hypoglycemia (glucose <3.5 mmol/L) in 6 of 15 participants after glibenclamide while there were no episodes of hypoglycemia after nateglinide. Among the case studies, nine individuals from six studies initiating SU or glinide therapy showed an average HbA1c decrement of 1.3% after an average of 12.2 months of treatment^30,33–35,39,43^ (Table 3).

We analyzed whether HNF1A-patients treated with insulin therapy can successfully transfer to SU. In a prospective study on HNF1A-diabetes (n=27) and HNF4A-diabetes (n=7) not on SU at diagnosis, 25 of the 31 patients on insulin discontinued insulin but only 12 (48%) achieved an HbA1c of 7.5%^22^. Good glycemia was associated with a shorter duration and lower HbA1c at transfer. An uncontrolled study of eight individuals with HNF1A-diabetes assessed success of transitioning from insulin that had been used since diabetes diagnosis (median insulin dose 0.5 units per kg; median duration of insulin treatment 20 years) to SU (gliclazide, median dose 80 mg daily)^28^ (Table 3). All patients were able to discontinue insulin. The HbA1c response was variable with a median reduction of 0.8% and six of eight improving but one individual had worsening of HbA1c by 3.2%. Table 3 also shows that seven individuals from five studies transitioning from insulin to SU showed an average HbA1c decrement of 1.35% after an average of 6.2 months of treatment with SU^32–33,37,41,43^.

Two randomized, double-blinded cross-over studies compared alternative and augmentative regimens to SU monotherapy in HNF1A-diabetes. Sixteen patients were enrolled in a trial comparing change in FPG and risk of hypoglycemia during 6 weeks of SU monotherapy (glimepiride) and 6 weeks of glucagon-like peptide-1 receptor agonist (GLP1RA) monotherapy (liraglutide) with 1-week washout between medications^26^. Among the 15 patients who completed the trial, FPG and post-prandial glucose was lowered by both treatments without a statistically significant difference between them. The number of mild hypoglycemic episodes (glucose <3.9 mmol/L) was markedly higher with glimepiride therapy (18 events) compared to liraglutide (1 event). The second trial compared the effects of 16 weeks of SU monotherapy (glimepiride) to 16 weeks of combination therapy with glimepiride and the dipeptidyl peptidase-4 inhibitor (DPP4i) linagliptin (with 4-week washout) on mean amplitude of glycemic excursions (MAGE) measured by CGM^27^. The combination therapy did not have an impact on MAGE over that of SU alone, but the mean (95% CI) HbA1c showed a significant decrease by −0.5% (−0.9 to −0.2, P = 0.0048) between SU and the combination therapy, included as a secondary end point.

There were only three studies where HNF4A data were reported separately^20,22,33^ (Table 3). Globa et al., report on two individuals with HNF4A-diabetes showing good response to SU, with >1% decrease in HbA1c (one was treatment-naive, the other switched from metformin)^33^. Another study included seven children with HNF4A-diabetes. Only two were on SU alone (mean HbA1c 7.0%, mean follow-up 8.3 years), while three were treated with insulin, two by choice and one due to inadequate control after switching from insulin to SU. Two others were not on pharmacologic treatment^20^.

#### 2. What is the optimal glucose lowering therapy in the two commonest subtypes of syndromic diabetes due to pathogenic variants in HNF1B and mitochondrial diabetes?

After duplicate removal and abstract screening of 1716 articles, 135 articles remained for full-text review. Data was extracted from 18 articles for HNF1B-diabetes, 42 for MD and 2 for both (Figure 2). We manually added an article not identified in the search^45^. There were no controlled trials, and the overall level of evidence was low and the risk of bias high. Tables 4 and 5 show a summary of the data for the 13 articles on HNF1B-diabetes^45–57^ and 10 articles on MD^45,58–66^ that reported any treatment response even in single cases, and the prevalent treatment modalities in patient cohorts. The articles represent altogether 293 cases with HNF1B-diabetes and 242 with m.3243A>G MD. Most articles only stated the current medication, which precludes drawing conclusions on the efficacy, but we note that at least one-fifth of the patients in both groups had no insulin treatment.

**Figure 2.**
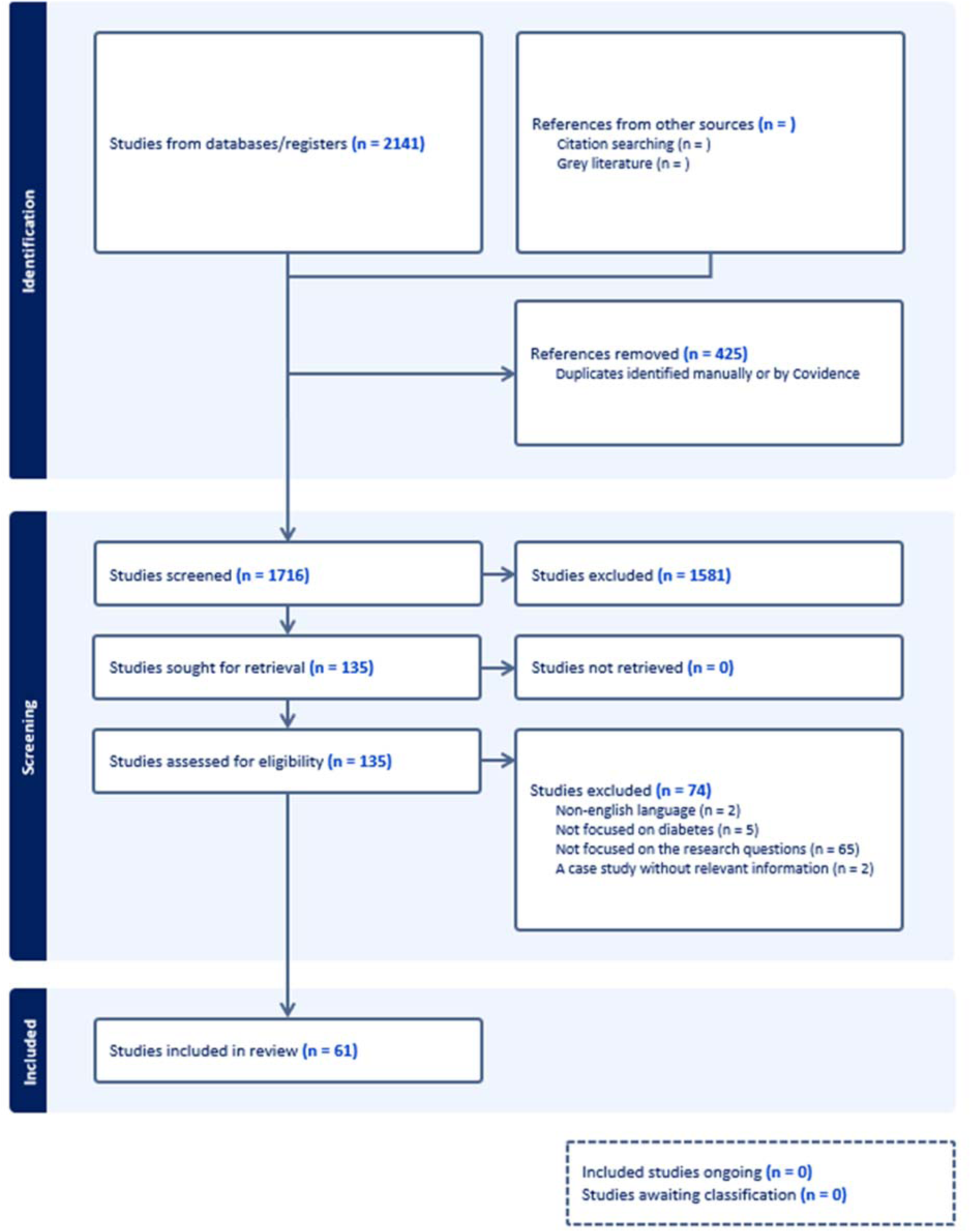
PRISMA search summary for HNF16-MODY and mitochondrial diabetes.

**Table 4.** Summary of included studies for HNF1B-diabetes.

| HNF1B-diabetes Cohort studies or case series (no randomized controlled trials) |  |  |  |  |  |  |  |  |  |  |
| --- | --- | --- | --- | --- | --- | --- | --- | --- | --- | --- |
| N=293 (132 men, 161 women) |  |  |  |  |  |  |  |  |  |  |
| Study ID | Number of participants | Sex (M/F) | Variant type (intra-genic/deletion) | Age (years) |  | Duration of diabetes (years) | Treatment before/after the genetic diagnosis |  | Intervention | Response |
|  |  |  |  | at diagnosis | at last assessment |  | before | after |  |  |
| Dubois-Laforgue 2017<br>*adult patients* | 159 | 73/86 | 75/84 | 28 [20–37] | 45 [3–56] | 12 [5.5–22.5] | 68 INS<br>47 OAD<br>25 LS# | 111 INS<br>22 OAD<br>7 LS |  |  |
| | 51* | | | | | 0.75 [0–5.25] | | | SU or repaglinide (response defined as HbA1c < 7%) | 29 responders (57%):<br>HbA1c<br>7.1% [5.5–12.1]<br>→ 6.1% [4.4–7.0]<br>($p < 10^{-4}$),<br>duration of<br>SU/repaglinide treatment<br>5 [3–9] years |
|  |  |  |  |  |  |  |  |  | INS replaced with SU in 10 patients | Three could convert |
| § Colclough 2022 | 50 | 26/24 | 26/24 | 18 [13–27] |  | 3 [1–8] | 23 INS<br>4<br>INS+OAD |  |  |  |
|  | 18 | 7/11 | 4/14 | 17 [12–25] |  | 3.5 [1–9] | 10 INS<br>4<br>INS+OAD |  |  |  |
| Warncke 2019<br>*children & adolescents* | 35 | 16/19 | NA (mostly deletions) | 13.5 [11.2–15.7] | 13.8 [12.4–16.3] | <1 | 20 INS<br>3<br>INS+OAD<br>4 OAD<br>8 LS |  |  |  |
| Ng 2022 | 10 | 6/4 | 9/1 | 31.5 [16–39] | 55 (18–62) | 18.5 (4–47) | 4 INS<br>2<br>INS+OAD<br>2 OAD<br>1 LS | 6 INS<br>2<br>INS+OAD<br>1 OAD (MET) | Trial of SU on 5 patients on INS | none successfully weaned off INS |
| Kettunen 2017 | 11 | 2/9 | 4/7 |  |  |  |  | 9 INS<br>2 LS |  |  |
| Case reports combined |  |  |  |  |  |  |  |  |  |  |
| † Roehlen 2018,<br>Terakawa 2020, | 10 | 2/8 | 5/5 | 16.5 [15.8–18.3] (N=8) | 22.5 [18.3–33.5] | 3 (0–22) | 5 INS<br>1 OAD (MET) | 3 INS<br>1<br>INS+OAD<br>2 OAD (MET) |  | Duration of non-INS response (HbA1c < 6.5–7%):<br>at least 1 year on GLP1RA (Terakawa 2020) |
| Carrillo 2017,<br>Tao 2020,<br>Aydın 2022, Ren 2021,<br>Thirumalai 2013,<br>Mateus 2020 |  |  |  |  |  |  |  | 1 GLPRA<br>1 LS |  | 6 years on SU+DPP4i<br>(Carrillo 2017)<br>at least 1 year on MET<br>(Tao 2020, Thirumalai 2013) |

Data are shown as median (range) or [interquartile range] or mean ± SD; M/F; M/F, number of male/female cases; NA, not available. INS, insulin; OAD, oral antidiabetic agent; SU, sulphonylurea; MET, metformin, GLP1RA, GLP1-receptor agonist; DPP4i, DPP4 inhibitor; TZD, thiazolidinedione; AGI, Alpha-glucosidase inhibitor
\* Those participating in the SU trial, included among the 159 patients above

##### a) HNF1B-diabetes

There were no randomized studies or trials of treatment in HNF1B-diabetes. Of the 168 individuals with HNF1B-diabetes, for whom treatment data after the genetic diagnosis was available, 132 (79%) used insulin, but it is not known if other medications had been tested (Table 4). One French study systematically evaluated the use of SU or repaglinide after the genetic diagnosis at a median [IQR] duration of 0.75 [0–5.25] years, and reported that 29 of 51 (57%) patients displayed an HbA1c decrease. However, the duration of SU or repaglinide treatment in those who did respond was 5 [3-9] years, and at a mean of 12 years follow-up, 79% of the cohort were on insulin^45^. They also tried replacing insulin with SU for 10 patients, which was successful in three (no details given). In an uncontrolled Irish study, none of five patients on insulin could successfully switch for sulfonylureas^48^.

##### b) Mitochondrial diabetes (MD) due to the m.3243A>G variant

There were no randomized studies or trials of treatment in m.3243 MD. Of the 233 individuals with MD, for whom treatment data after the genetic diagnosis was available, 167 (72%) used insulin, but again it is not known if other medications had been tested (Table 5). No studies are reported trying to discontinue insulin by treating with OHAs.

**Table 5.**
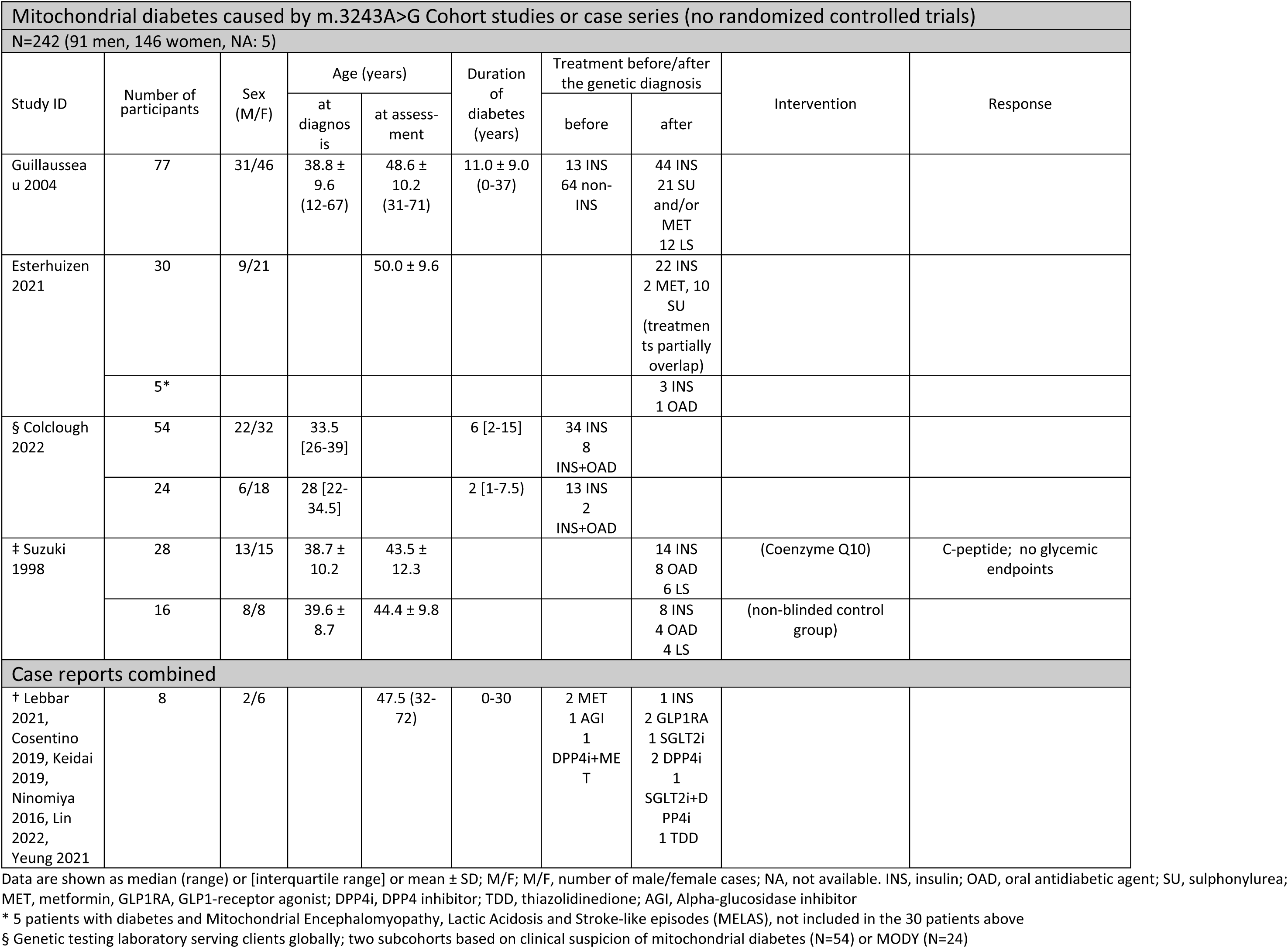
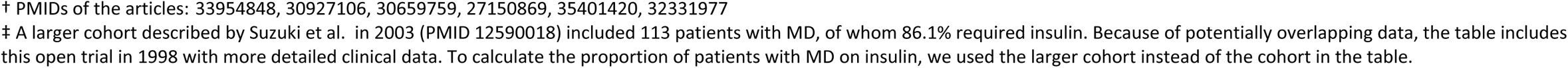
Summary of included studies for MD.

| Mitochondrial diabetes caused by m.3243A>G Cohort studies or case series (no randomized controlled trials) |  |  |  |  |  |  |  |  |  |
| --- | --- | --- | --- | --- | --- | --- | --- | --- | --- |
| N=242 (91 men, 146 women, NA: 5) |  |  |  |  |  |  |  |  |  |
| Study ID | Number of participants | Sex (M/F) | Age (years) |  | Duration of diabetes (years) | Treatment before/after the genetic diagnosis |  | Intervention | Response |
|  |  |  | at diagnosis | at assessment |  | before | after |  |  |
| Guillausseau 2004 | 77 | 31/46 | 38.8 ± 9.6 (12-67) | 48.6 ± 10.2 (31-71) | 11.0 ± 9.0 (0-37) | 13 INS<br>64 non-INS | 44 INS<br>21 SU and/or MET<br>12 LS |  |  |
| Esterhuizen 2021 | 30 | 9/21 |  | 50.0 ± 9.6 |  |  | 22 INS<br>2 MET, 10 SU<br>(treatments partially overlap) |  |  |
|  | 5* |  |  |  |  |  | 3 INS<br>1 OAD |  |  |
| § Colclough 2022 | 54 | 22/32 | 33.5 [26-39] |  | 6 [2-15] | 34 INS<br>8 INS+OAD |  |  |  |
|  | 24 | 6/18 | 28 [22-34.5] |  | 2 [1-7.5] | 13 INS<br>2 INS+OAD |  |  |  |
| ‡ Suzuki 1998 | 28 | 13/15 | 38.7 ± 10.2 | 43.5 ± 12.3 |  |  | 14 INS<br>8 OAD<br>6 LS | (Coenzyme Q10) | C-peptide; no glycemic endpoints |
|  | 16 | 8/8 | 39.6 ± 8.7 | 44.4 ± 9.8 |  |  | 8 INS<br>4 OAD<br>4 LS | (non-blinded control group) |  |
| Case reports combined |  |  |  |  |  |  |  |  |  |
| † Lebbar 2021, Cosentino 2019, Keidai 2019, Ninomiya 2016, Lin 2022, Yeung 2021 | 8 | 2/6 |  | 47.5 (32-72) | 0-30 | 2 MET<br>1 AGI<br>1 DPP4i+MET | 1 INS<br>2 GLP1RA<br>1 SGLT2i<br>2 DPP4i<br>1 SGLT2i+DPP4i<br>1 TDD |  |  |

Data are shown as median (range) or [interquartile range] or mean ± SD; M/F; M/F, number of male/female cases; NA, not available. INS, insulin; OAD, oral antidiabetic agent; SU, sulphonylurea; MET, metformin, GLP1RA, GLP1-receptor agonist; DPP4i, DPP4 inhibitor; TDD, thiazolidinedione; AGI, Alpha-glucosidase inhibitor
\* 5 patients with diabetes and Mitochondrial Encephalomyopathy, Lactic Acidosis and Stroke-like episodes (MELAS), not included in the 30 patients above
§ Genetic testing laboratory serving clients globally; two subcohorts based on clinical suspicion of mitochondrial diabetes (N=54) or MODY (N=24)
† PMIDs of the articles: 33954848, 30927106, 30659759, 27150869, 35401420, 32331977
‡ A larger cohort described by Suzuki et al. in 2003 (PMID 12590018) included 113 patients with MD, of whom 86.1% required insulin. Because of potentially overlapping data, the table includes this open trial in 1998 with more detailed clinical data. To calculate the proportion of patients with MD on insulin, we used the larger cohort instead of the cohort in the table.

Evidence against the use of metformin was limited to three case reports^65,67–68^. In two of them, metformin use was associated with elevated lactate levels (2.5-3.7 mmol/L^67^, and up to 5.9 mmol/L^65^), and the first was reported to have lactic acidosis. However, pH was not given for either case, and an improved lactate level (2.4 mmol/L) after discontinuation of metformin was only given for the latter case.

#### 3. Are there alternatives to insulin therapy in 6q24 transient neonatal diabetes (6q24-TND) and in SLC19A2-diabetes, also known as Thiamine-Responsive Megaloblastic Anemia Syndrome (TRMA) does thiamine supplementation improve glycemia?

##### a) 6q24-TND

The literature search identified 1489 studies related to 6q24 TNMD (Figure 3). After duplicate removal, abstract screening, and full-text review, 19 studies met eligibility criteria, including five case series reporting on 16 cases, 14 reports of single cases, for a total of 30 6q24 cases for whom data was available regarding treatment with non-insulin therapies (Table 6).

**Figure 3.**
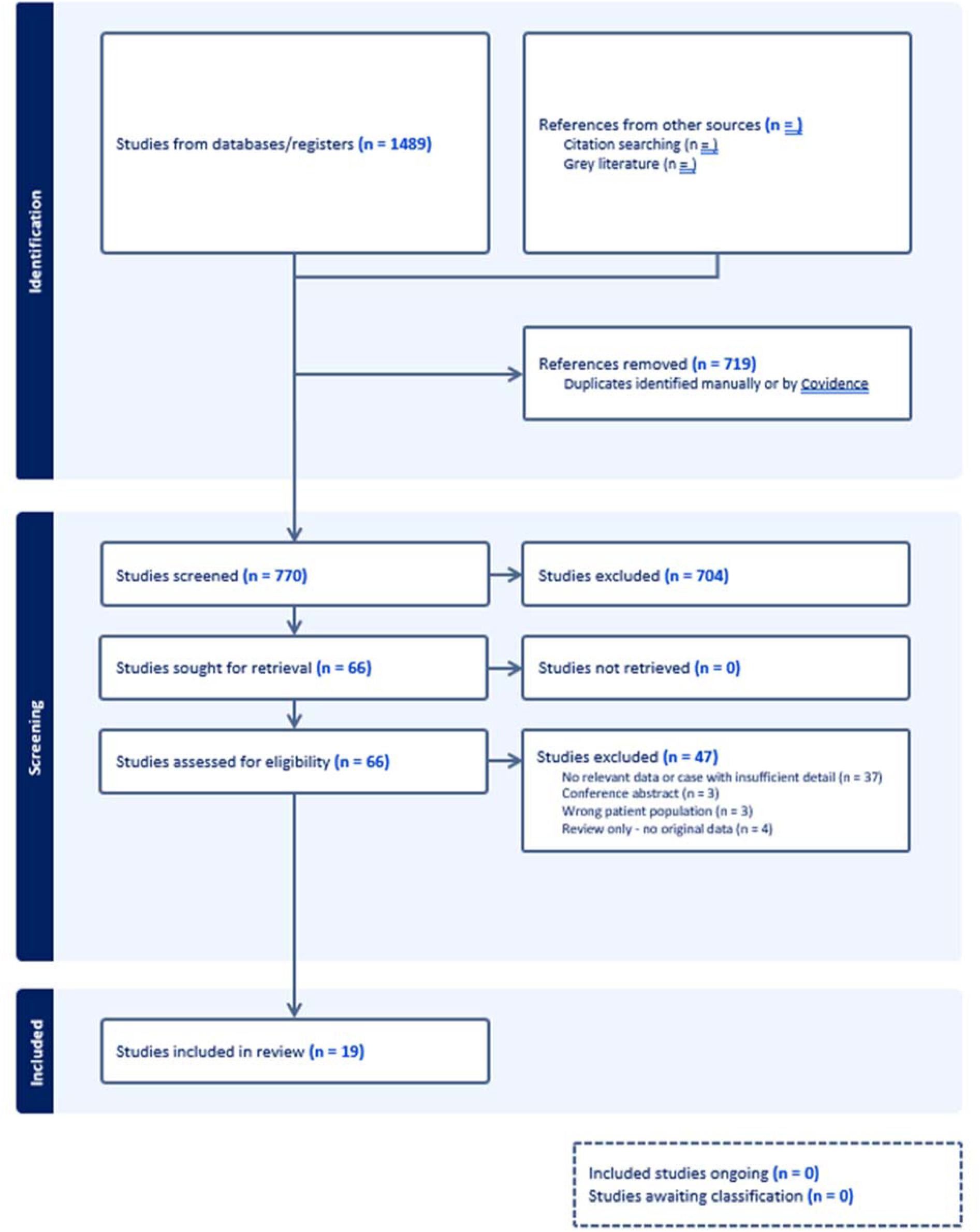
PRISMA search summary for 6q24-related transient neonatal diabetes.

**Table 6.** Summary of included studies for 6q24 TND.

| 6q24 Case reports with use of non-insulin therapies during neonatal phase |  |  |  |  |  |  |  |  |  |  |  |  |
| --- | --- | --- | --- | --- | --- | --- | --- | --- | --- | --- | --- | --- |
| Study ID | 6q24 Mechanism (UPD6, PD, MD, NR) | Sex | Birth weight (grams) | Age at Dx of DM (days) | SU successful? | Days until insulin stopped |  | Maximum SU dose (mg/kg/d) | (G | Age at remission of diabetes (weeks) |  |  |
| Gore 2020 | PD | F | 1820 | 1 | Yes | 5 |  | 1 |  | 7 |  |  |
| Neumann 2018 | UPD6 | M | 1700 | 1 | Yes | 26 |  | 1.2 |  | 16 |  |  |
| Cao 2017 | UPD6 | F | 2000 | 1 | No | NA |  | NR |  | NA* |  |  |
| Zhang 2015 | MD | F | 1740 | 24 | Yes | 5 |  | 0.09 |  | NR |  |  |
| Yao 2014 | UPD6 | F | 2000 | 2 | No | 7 |  | 0.1 |  | NA* |  |  |
| Hewes 2010 | NR | M | 2720 | 8 | No | NA |  | NR |  | 12 |  |  |
| Senguttuvan 2015 | PD | M | NR | 2 | No | NA |  | NR |  | 12 |  |  |
| Carmody 2014 | NR | NR | NR | 14 | Yes | 14 |  | NR |  | 6 |  |  |
|  | NR | NR | NR | 4 | Yes | 11 |  | NR |  | 4 |  |  |
|  | NR | NR | NR | 1 | Yes | 1 |  | NR |  | 14 |  |  |
| Li 2018 | UPD6 | NR | NR | 25 | Yes | NR |  | 0.52 |  | 16 |  |  |
|  | UPD6 | NR | NR | 4 | Yes | 13 |  | 0.4 |  | 12 |  |  |
|  | PD | NR | NR | 8 | No | NA |  | NR |  | 30 |  |  |
|  | UPD6 | NR | NR | 2 | No | NA |  | NR |  | NA* |  |  |
| Garcin 2018 | UPD6 | M | 1370 | 1 | Yes | 3 |  | 0.15 |  | 18 |  |  |
|  | PD | F | NR | 1 | No | NA |  | 4 |  | NR |  |  |
| 6q24 Case reports with use of non-insulin therapies during relapse phase |  |  |  |  |  |  |  |  |  |  |  |  |
| Study ID | 6q24 Mechanism (UPD6, PD, MD, NR) | Sex | Birth weight (grams) | Age at Dx of DM (days) | Age at remission of neonatal DM (weeks) | Age of DM Relapse (years) | Insulin dose prior to NIT (U/kg/d) | Age at start of NIT (years) | NIT Used | Insulin discontinued? Y/N | HbA1c before NIT | HbA1c after NIT |
| Schimmel 2009 | UPD6 | M | 2060 | 2 | 28 | 15 | 0 | 15 | Glim | Yes** | NR | 5.7-7.8 |
| Kontbay 2022 | MD | F | 3000 | 1 | 12 | 8.5 | 0.25 | 8.5 | Glib | Yes | NR | 8.7 |
| Sato 2021 | PD | M | 2226 | 2 | 26 | 14.5 | 0 | 16 | Glib | Yes** | 6.7 | NR |
| Fu 2019 | MD | M | 1700 | 45 | 12 | 14 | 0.2 | 14 | Glim | Yes | 7.8 | 6.5-7 |
| Yorifuji 2014 | PD | M | 1660 | 7 | 28 | 12 | 0 | 12 | Vog, Alo | Yes** | 7.5 | 6.2 |
| vondemBerge 2021 | MD | NR | NR | 8 | 6 | 14 | 0.7 | 14 | Glib | Yes | 8.2 | 5.7 |
| Uchida 2020 | PD | F | 1765 | 11 | 8 | 10.8 | 0 | 10.8 | Met | Yes | 7.4 | 6.5 |
| Garcin 2018 | UPD6 | F | NR | 1 | 32 | 12 | 0 | 12 | Glib | Yes** | NR | NR |
| Sovik 2012 | PD | M | 1840 | 6 | 12 | Adult | 0 | 43 | Met, Glib | Yes** | NR | 9.0 |
|  | PD | F | 1440 | 12 | 54 | 11 | 0.5 | 28 | Met | No | 8.8 | 10.0 |
| Carmody 2015 | UPD6 | F | 1280 | 1 | 16 | 13 | 0.6 | 20 | Glib, Sit, Met | Yes | 82 | 7.1 |
|  | UPD6 | M | 2470 | 1 | 28 | 12 | 0.7 | 23 | Glib, Sit, Met | Yes | 7.8 | 6.6 |
|  | UPD6 | F | 2240 | 1 | 24 | 27 | 0.4 | 29 | Glib | Yes | 7.2 | 7.3 |
|  | UPD6 | F | 1810 | 1 | 12 | 12 | 0.8 | 28 | Glib | Yes | 9.9 | 7.5 |

NR: not reported; NA: not applicable; G: glibenclamide; UPD6: uniparental paternal disomy of chromosome 6; PD: paternally-inherited duplication involving 6q24; MD: maternal methylation defect (either ZFP57 mutations or unknown cause); NA\*: continued to require insulin as old as 41-60 months.
NIT: non-insulin therapy; Yes\*\*: insulin never given during relapse phase; Glib: glibenclamide (glyburide); Glim: glimepiride; Vog: voglibose (alpha-glucosidase inhibitor); Alo: alogliptin (DPP4-inhibitor); Met: metformin.

There were no randomized trials for therapeutic response in 6q24 TNDM. For 16 cases with relevant data on the initial neonatal phase of diabetes^69–78^, SU (glyburide or glibenclamide in nearly all cases) was the only class of medication used other than insulin. Efficacy of SU during the neonatal phase was inconsistent, with studies reporting no effect or failure of SU to improve diabetes management in seven cases^71,73–75,77–78^, while for nine cases SU treatment was reported to allow insulin to be discontinued^69–70,72,76–78^. Of note, in most cases the diabetes remitted within days to weeks after insulin was discontinued, but three cases were reported to remain insulin-treated as old as 41-60 months of age at the time of the reports^71,73,77^. None of these three cases with a possibly more permanent neonatal diabetes phenotype exhibited a response to SU treatment.

For case reports with relevant data on the use of non-insulin therapies during the later relapse phase of diabetes^78–87^, apparent efficacy of such treatment was more consistent, with 13 of 14 cases being either able to discontinue insulin at least temporarily or were never started on insulin (three cases)^78–85,87^ (Table 6). There was a wide range of follow-up time after the initiation of non-insulin therapies, but in most cases it was at least 6 months, and in a few cases many years. Measures of glycemia were reported inconsistently. The most common class of medications utilized was SU (most commonly glyburide or glibenclamide), with some reporting use of metformin (either as monotherapy or as an adjunctive agent), and few cases utilized DPP4i (either alone or with SU). For the one patient who was not able to discontinue insulin, metformin was the only additional agent utilized^86^. The only adverse events reported were rare mild gastrointestinal symptoms and mild hypoglycemia, but these may have been under reported.

##### b) SLC19A2-diabetes

The literature search yielded 166 studies (Figure 4). After duplicate removal, abstract screening, and full-text review, 32 studies were included in the review with three larger case series and 29 case reports. Data with varying extent of outcome measures on the effect of thiamine therapy on SLC19A2-diabetes were available for 95 patients, with data at the individual level for 72 patients, and at group level for 23 patients in one case series (Tables 7 and 8).

**Figure 4.**
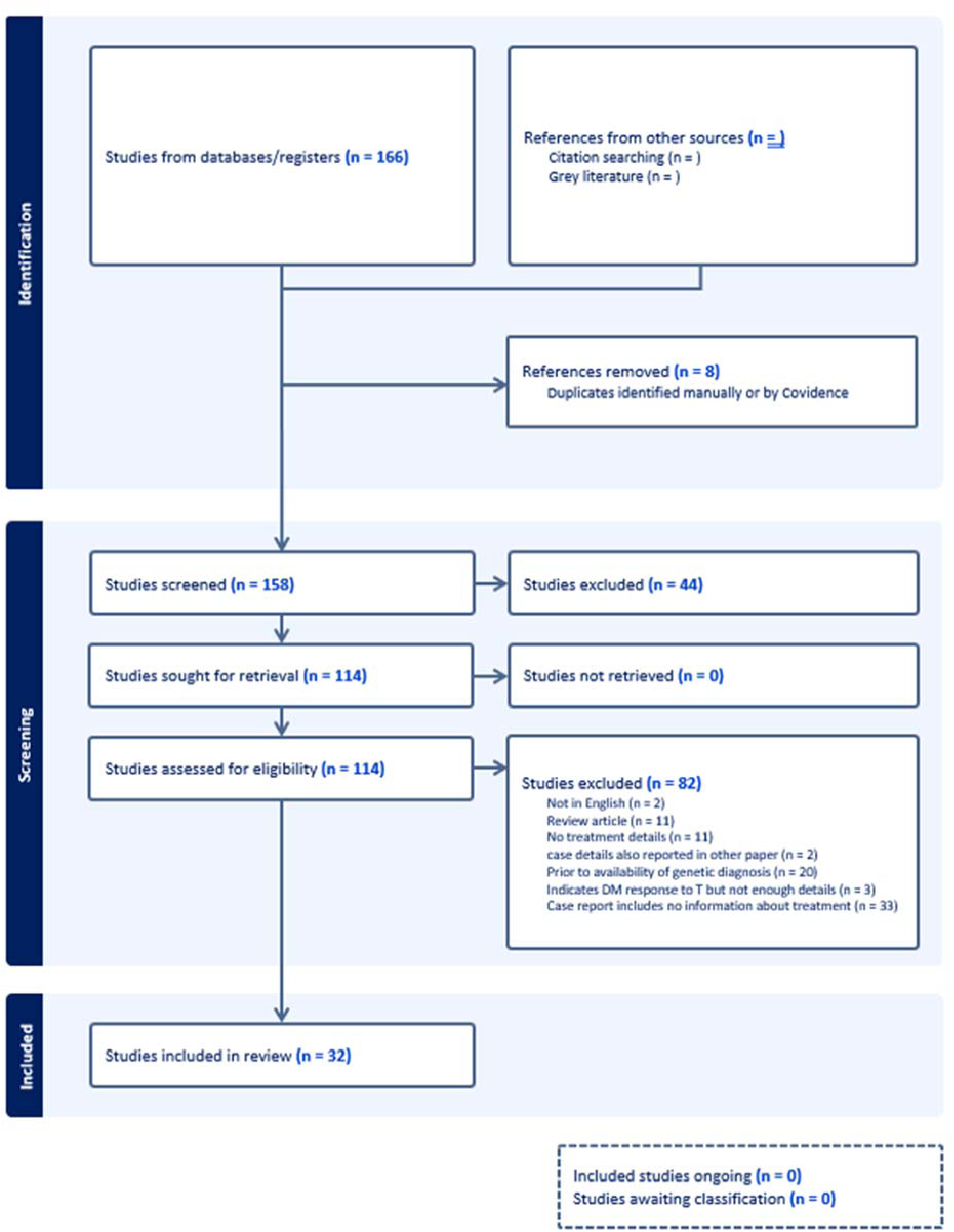
PRI MA search summary for SLC19A2-diabetes.

**Table 7.**
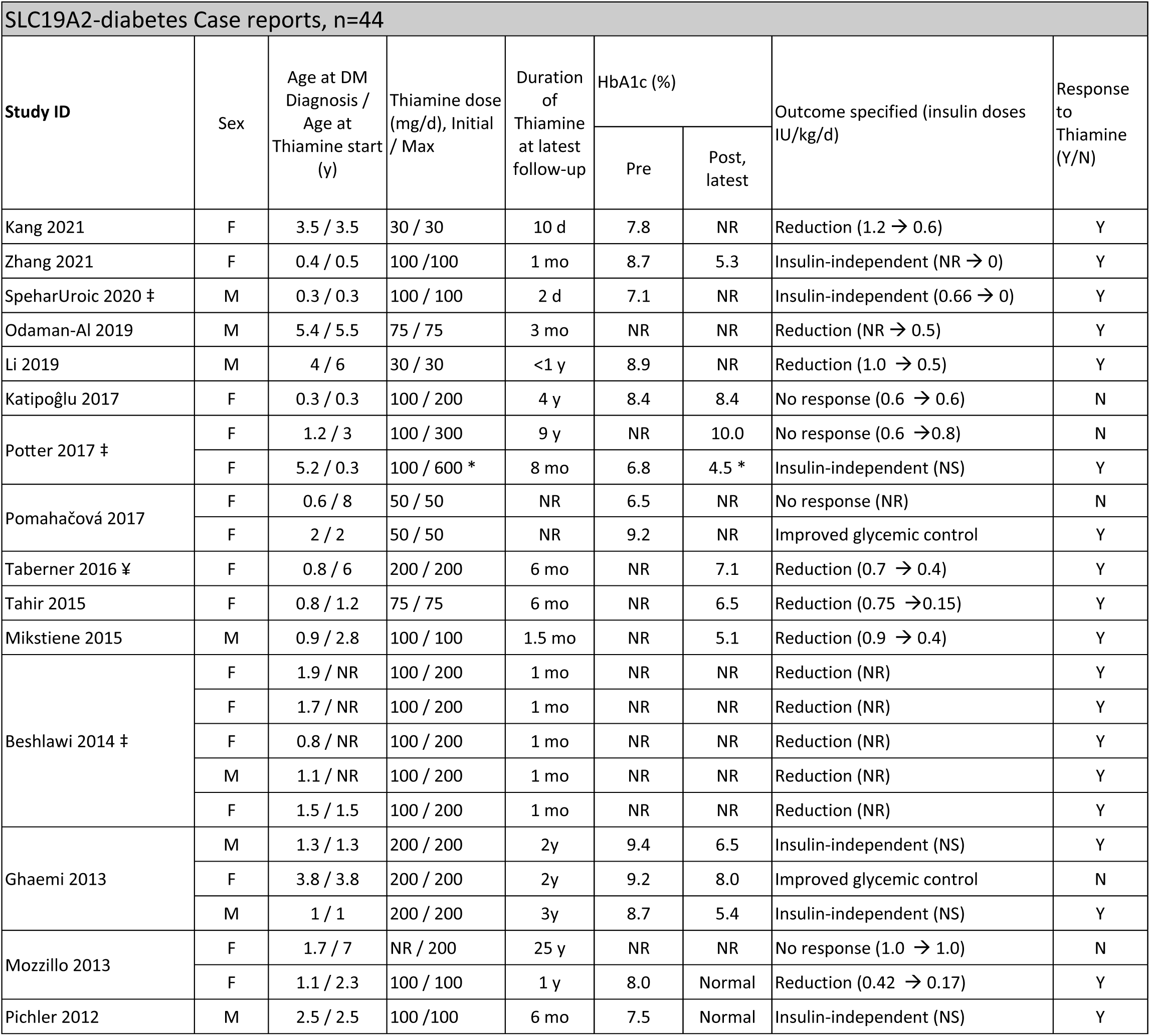
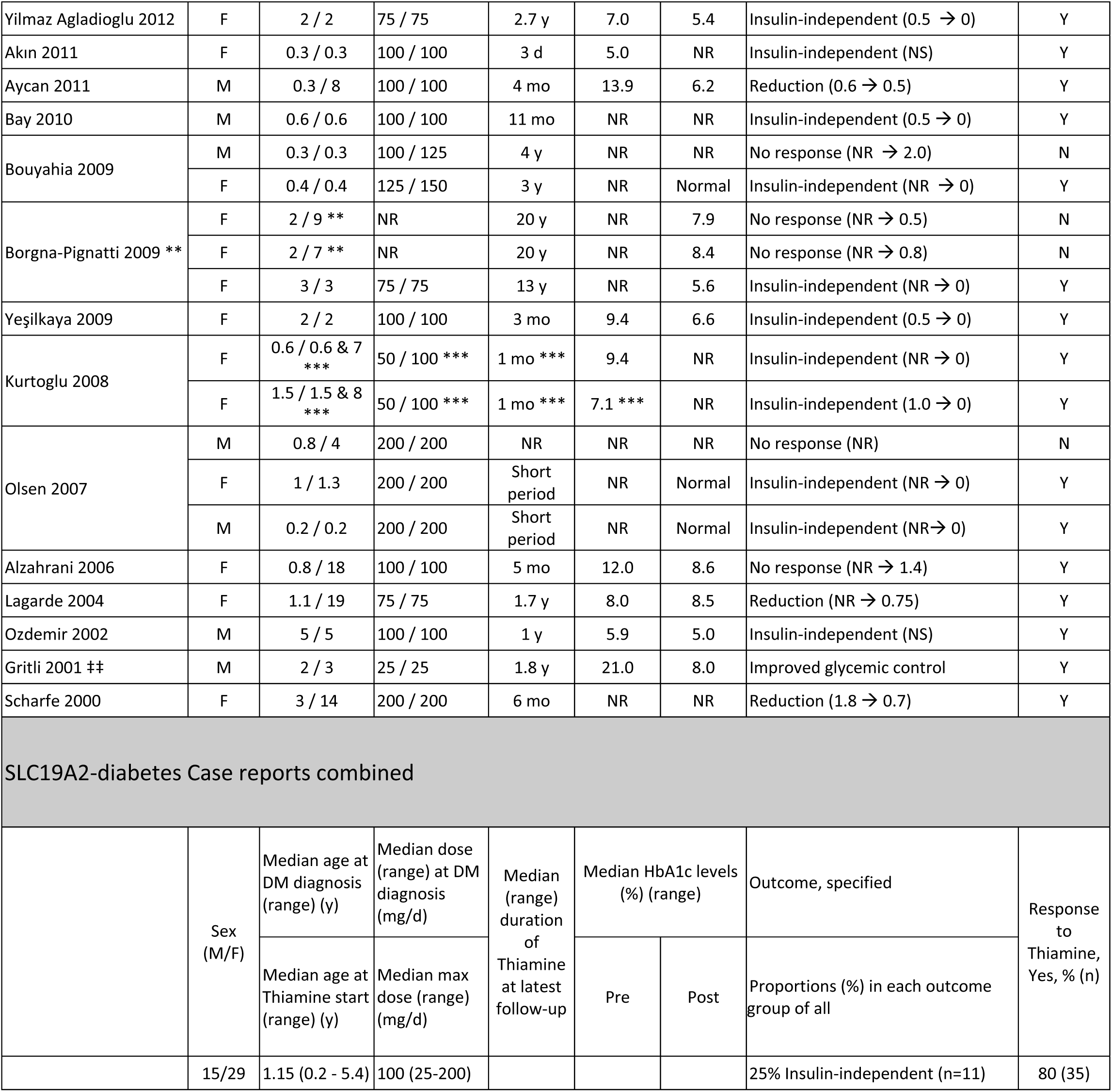
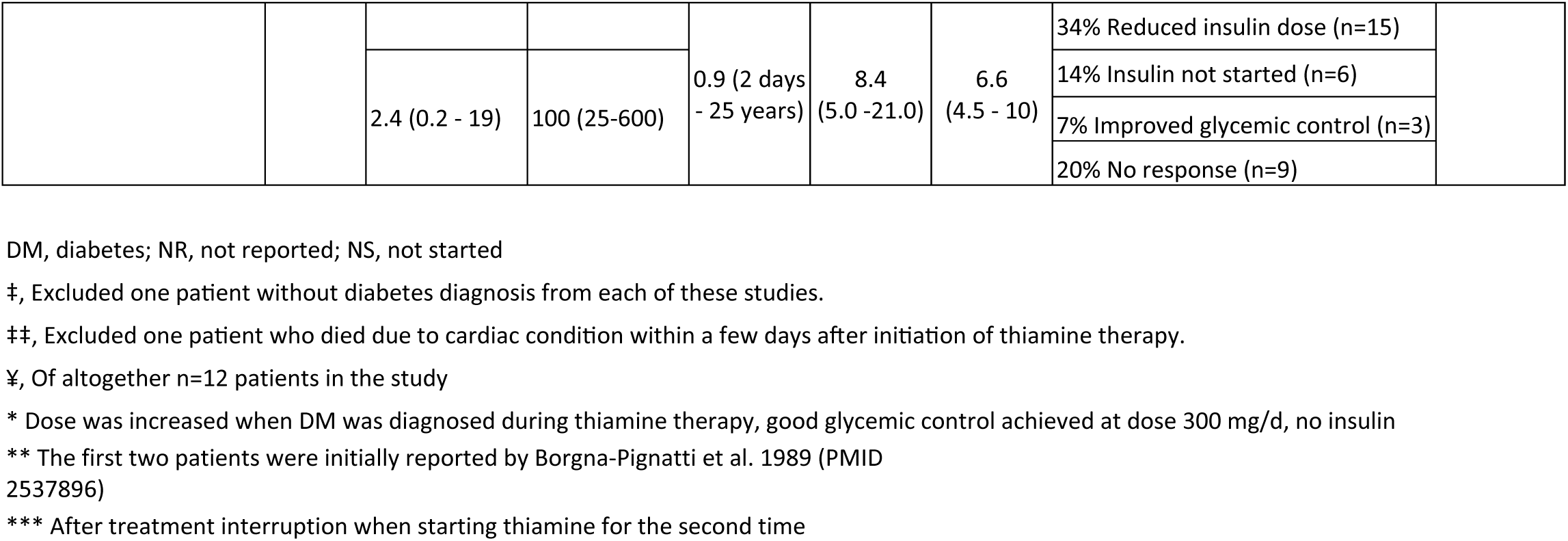
Summary of included studies for SLC19A2-diabetes (TRMA syndrome.

| SLC19A2-diabetes Case reports, n=44 |  |  |  |  |  |  |  |  |
| --- | --- | --- | --- | --- | --- | --- | --- | --- |
| Study ID | Sex | Age at DM Diagnosis / Age at Thiamine start (y) | Thiamine dose (mg/d), Initial / Max | Duration of Thiamine at latest follow-up | HbA1c (%) |  | Outcome specified (insulin doses IU/kg/d) | Response to Thiamine (Y/N) |
|  |  |  |  |  | Pre | Post, latest |  |  |
| Kang 2021 | F | 3.5 / 3.5 | 30 / 30 | 10 d | 7.8 | NR | Reduction (1.2 → 0.6) | Y |
| Zhang 2021 | F | 0.4 / 0.5 | 100 / 100 | 1 mo | 8.7 | 5.3 | Insulin-independent (NR → 0) | Y |
| SpeharUroic 2020 ‡ | M | 0.3 / 0.3 | 100 / 100 | 2 d | 7.1 | NR | Insulin-independent (0.66 → 0) | Y |
| Odaman-AI 2019 | M | 5.4 / 5.5 | 75 / 75 | 3 mo | NR | NR | Reduction (NR → 0.5) | Y |
| Li 2019 | M | 4 / 6 | 30 / 30 | <1 y | 8.9 | NR | Reduction (1.0 → 0.5) | Y |
| Katipoğlu 2017 | F | 0.3 / 0.3 | 100 / 200 | 4 y | 8.4 | 8.4 | No response (0.6 → 0.6) | N |
| Potter 2017 ‡ | F | 1.2 / 3 | 100 / 300 | 9 y | NR | 10.0 | No response (0.6 → 0.8) | N |
|  | F | 5.2 / 0.3 | 100 / 600 * | 8 mo | 6.8 | 4.5 * | Insulin-independent (NS) | Y |
| Pomahačová 2017 | F | 0.6 / 8 | 50 / 50 | NR | 6.5 | NR | No response (NR) | N |
|  | F | 2 / 2 | 50 / 50 | NR | 9.2 | NR | Improved glycemic control | Y |
| Taberner 2016 ¥ | F | 0.8 / 6 | 200 / 200 | 6 mo | NR | 7.1 | Reduction (0.7 → 0.4) | Y |
| Tahir 2015 | F | 0.8 / 1.2 | 75 / 75 | 6 mo | NR | 6.5 | Reduction (0.75 → 0.15) | Y |
| Mikstiene 2015 | M | 0.9 / 2.8 | 100 / 100 | 1.5 mo | NR | 5.1 | Reduction (0.9 → 0.4) | Y |
| Beshlawi 2014 ‡ | F | 1.9 / NR | 100 / 200 | 1 mo | NR | NR | Reduction (NR) | Y |
|  | F | 1.7 / NR | 100 / 200 | 1 mo | NR | NR | Reduction (NR) | Y |
|  | F | 0.8 / NR | 100 / 200 | 1 mo | NR | NR | Reduction (NR) | Y |
|  | M | 1.1 / NR | 100 / 200 | 1 mo | NR | NR | Reduction (NR) | Y |
|  | F | 1.5 / 1.5 | 100 / 200 | 1 mo | NR | NR | Reduction (NR) | Y |
| Ghaemi 2013 | M | 1.3 / 1.3 | 200 / 200 | 2y | 9.4 | 6.5 | Insulin-independent (NS) | Y |
|  | F | 3.8 / 3.8 | 200 / 200 | 2y | 9.2 | 8.0 | Improved glycemic control | N |
|  | M | 1 / 1 | 200 / 200 | 3y | 8.7 | 5.4 | Insulin-independent (NS) | Y |
| Mozzillo 2013 | F | 1.7 / 7 | NR / 200 | 25 y | NR | NR | No response (1.0 → 1.0) | N |
|  | F | 1.1 / 2.3 | 100 / 100 | 1 y | 8.0 | Normal | Reduction (0.42 → 0.17) | Y |
| Pichler 2012 | M | 2.5 / 2.5 | 100 / 100 | 6 mo | 7.5 | Normal | Insulin-independent (NS) | Y |
| Yilmaz Agladioglu 2012 | F | 2 / 2 | 75 / 75 | 2.7 y | 7.0 | 5.4 | Insulin-independent (0.5 → 0) | Y |
| Akın 2011 | F | 0.3 / 0.3 | 100 / 100 | 3 d | 5.0 | NR | Insulin-independent (NS) | Y |
| Ayçan 2011 | M | 0.3 / 8 | 100 / 100 | 4 mo | 13.9 | 6.2 | Reduction (0.6 → 0.5) | Y |
| Bay 2010 | M | 0.6 / 0.6 | 100 / 100 | 11 mo | NR | NR | Insulin-independent (0.5 → 0) | Y |
| Bouyahia 2009 | M | 0.3 / 0.3 | 100 / 125 | 4 y | NR | NR | No response (NR → 2.0) | N |
|  | F | 0.4 / 0.4 | 125 / 150 | 3 y | NR | Normal | Insulin-independent (NR → 0) | Y |
| Borgna-Pignatti 2009 ** | F | 2 / 9 ** | NR | 20 y | NR | 7.9 | No response (NR → 0.5) | N |
|  | F | 2 / 7 ** | NR | 20 y | NR | 8.4 | No response (NR → 0.8) | N |
|  | F | 3 / 3 | 75 / 75 | 13 y | NR | 5.6 | Insulin-independent (NR → 0) | Y |
| Yeşilkaya 2009 | F | 2 / 2 | 100 / 100 | 3 mo | 9.4 | 6.6 | Insulin-independent (0.5 → 0) | Y |
| Kurtoglu 2008 | F | 0.6 / 0.6 & 7 *** | 50 / 100 *** | 1 mo *** | 9.4 | NR | Insulin-independent (NR → 0) | Y |
|  | F | 1.5 / 1.5 & 8 *** | 50 / 100 *** | 1 mo *** | 7.1 *** | NR | Insulin-independent (1.0 → 0) | Y |
| Olsen 2007 | M | 0.8 / 4 | 200 / 200 | NR | NR | NR | No response (NR) | N |
|  | F | 1 / 1.3 | 200 / 200 | Short period | NR | Normal | Insulin-independent (NR → 0) | Y |
|  | M | 0.2 / 0.2 | 200 / 200 | Short period | NR | Normal | Insulin-independent (NR → 0) | Y |
| Alzahrani 2006 | F | 0.8 / 18 | 100 / 100 | 5 mo | 12.0 | 8.6 | No response (NR → 1.4) | Y |
| Lagarde 2004 | F | 1.1 / 19 | 75 / 75 | 1.7 y | 8.0 | 8.5 | Reduction (NR → 0.75) | Y |
| Ozdemir 2002 | M | 5 / 5 | 100 / 100 | 1 y | 5.9 | 5.0 | Insulin-independent (NS) | Y |
| Gritli 2001 †† | M | 2 / 3 | 25 / 25 | 1.8 y | 21.0 | 8.0 | Improved glycemic control | Y |
| Scharfe 2000 | F | 3 / 14 | 200 / 200 | 6 mo | NR | NR | Reduction (1.8 → 0.7) | Y |

SLC19A2-diabetes Case reports combined
|  | Sex<br>(M/F) | Median age at<br>DM diagnosis<br>(range) (y) | Median dose<br>(range) at DM<br>diagnosis<br>(mg/d) | Median<br>(range)<br>duration<br>of<br>Thiamine<br>at latest<br>follow-up | Median HbA1c levels<br>(%) (range) |  | Outcome, specified | Response<br>to<br>Thiamine,<br>Yes, % (n) |
| --- | --- | --- | --- | --- | --- | --- | --- | --- |
|  |  | Median age at<br>Thiamine start<br>(range) (y) | Median max<br>dose (range)<br>(mg/d) |  | Pre | Post | Proportions (%) in each outcome<br>group of all |  |
|  | 15/29 | 1.15 (0.2 - 5.4) | 100 (25-200) |  |  |  | 25% Insulin-independent (n=11) | 80 (35) |

|  |  |  |  |  |  |  |  |
| --- | --- | --- | --- | --- | --- | --- | --- |
|  |  |  |  | 0.9 (2 days<br>- 25 years) | 8.4<br>(5.0 -21.0) | 6.6<br>(4.5 - 10) | 34% Reduced insulin dose (n=15) |
|  |  | 2.4 (0.2 - 19) | 100 (25-600) |  |  |  | 14% Insulin not started (n=6) |
|  |  |  |  |  |  |  | 7% Improved glycemic control (n=3) |
|  |  |  |  |  |  |  | 20% No response (n=9) |

DM, diabetes; NR, not reported; NS, not started
‡, Excluded one patient without diabetes diagnosis from each of these studies.
‡‡, Excluded one patient who died due to cardiac condition within a few days after initiation of thiamine therapy.
¥, Of altogether n=12 patients in the study
\* Dose was increased when DM was diagnosed during thiamine therapy, good glycemic control achieved at dose 300 mg/d, no insulin
\*\* The first two patients were initially reported by Borgna-Pignatti et al. 1989 (PMID 2537896)
\*\*\* After treatment interruption when starting thiamine for the second time

**Table 8.**
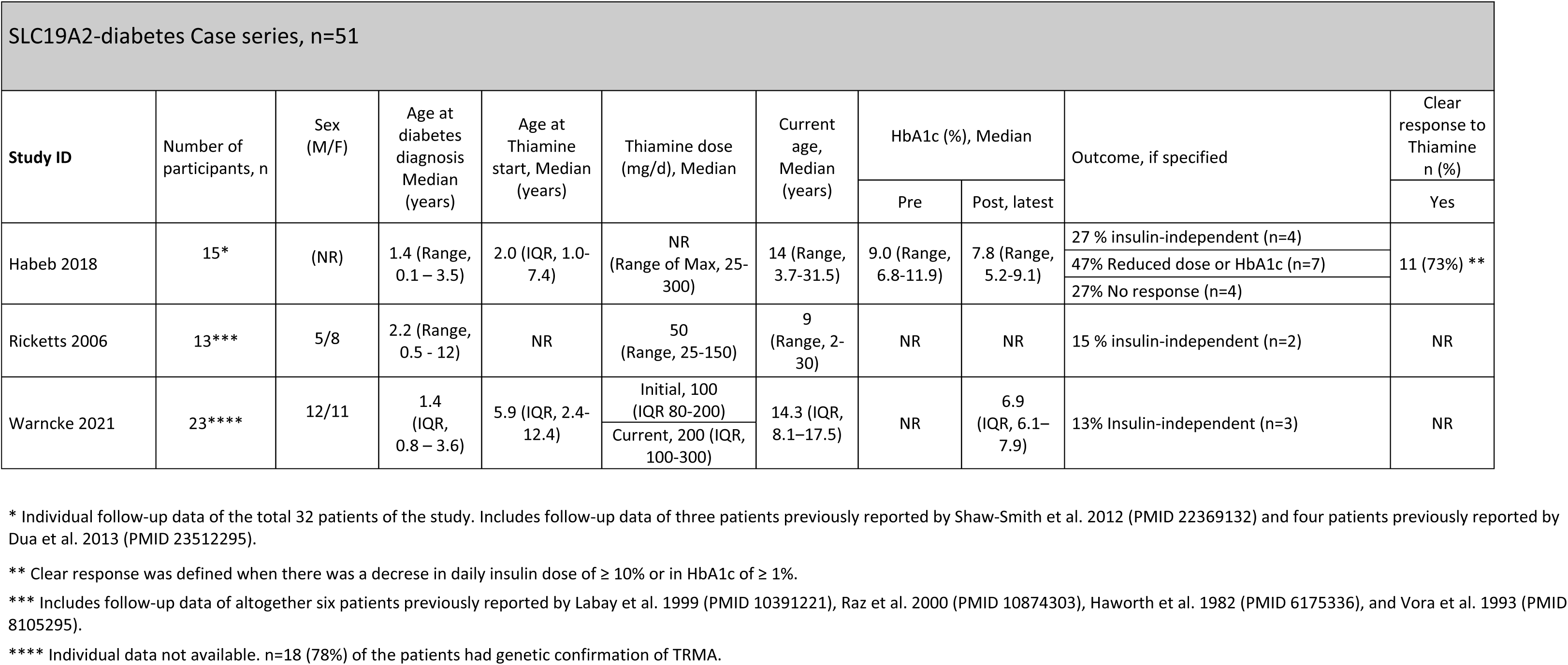
Summary of included studies for SLC19A2-diabetes (TRMA syndrome)

| SLC19A2-diabetes Case series, n=51 |  |  |  |  |  |  |  |  |  |  |
| --- | --- | --- | --- | --- | --- | --- | --- | --- | --- | --- |
| Study ID | Number of participants, n | Sex (M/F) | Age at diabetes diagnosis Median (years) | Age at Thiamine start, Median (years) | Thiamine dose (mg/d), Median | Current age, Median (years) | HbA1c (%), Median |  | Outcome, if specified | Clear response to Thiamine n (%) |
|  |  |  |  |  |  |  | Pre | Post, latest |  | Yes |
| Habeb 2018 | 15* | (NR) | 1.4 (Range, 0.1 – 3.5) | 2.0 (IQR, 1.0-7.4) | NR (Range of Max, 25-300) | 14 (Range, 3.7-31.5) | 9.0 (Range, 6.8-11.9) | 7.8 (Range, 5.2-9.1) | 27 % insulin-independent (n=4) | 11 (73%) ** |
|  |  |  |  |  |  |  |  |  | 47% Reduced dose or HbA1c (n=7) |  |
|  |  |  |  |  |  |  |  |  | 27% No response (n=4) |  |
| Ricketts 2006 | 13*** | 5/8 | 2.2 (Range, 0.5 - 12) | NR | 50 (Range, 25-150) | 9 (Range, 2-30) | NR | NR | 15 % insulin-independent (n=2) | NR |
| Warncke 2021 | 23**** | 12/11 | 1.4 (IQR, 0.8 – 3.6) | 5.9 (IQR, 2.4-12.4) | Initial, 100 (IQR 80-200) | 14.3 (IQR, 8.1–17.5) | NR | 6.9 (IQR, 6.1–7.9) | 13% Insulin-independent (n=3) | NR |
|  |  |  |  |  | Current, 200 (IQR, 100-300) |  |  |  |  |  |

\* Individual follow-up data of the total 32 patients of the study. Includes follow-up data of three patients previously reported by Shaw-Smith et al. 2012 (PMID 22369132) and four patients previously reported by Dua et al. 2013 (PMID 23512295).
\*\* Clear response was defined when there was a decrease in daily insulin dose of $\geq 10\%$ or in HbA1c of $\geq 1\%$ .
\*\*\* Includes follow-up data of altogether six patients previously reported by Labay et al. 1999 (PMID 10391221), Raz et al. 2000 (PMID 10874303), Haworth et al. 1982 (PMID 6175336), and Vora et al. 1993 (PMID 8105295).
\*\*\*\* Individual data not available. n=18 (78%) of the patients had genetic confirmation of TRMA.

There were no randomized trials for therapeutic response in SLC19A2-diabetes, and the scope of the studies was mainly on overall description of the phenotype.

The data from case reports and case series are shown in tables 7 and 8, respectively. Diabetes was diagnosed at the median age of 1.15 (range, 0.2 – 5.4, n=44) years in case reports^88–116^ and between median ages of 1.4 and 2.2 (0.1 – 12, n=51) years in the three case series^117–119^. Insulin was the most common therapy for diabetes (n=89/95) with one patient also using SU (glimepiride), and 6 not receiving any antidiabetic therapy. Thiamine therapy was initiated after a median duration of diabetes of 4 months (0 – 17.9 years, n=53/95), two additional patients were already on thiamine therapy at the time of diabetes onset.

The median thiamine treatment duration at the time of follow-up was 0.9 years (3 days – 25 years, n=39) in case reports, 4.7 years (2–17.5 years, n=15) in one case series, and not available in the two remaining case series. Effect of thiamine treatment on glycemic control was inconsistently reported using different outcome measures. Regarding the 72 patients with individual data, commencement of thiamine treatment was associated with: achievement of insulin-independency in 24% (n=17), decrease in daily insulin requirement in 38% (n=27), improvement in glycemic control without specification in 4% (n=3), no response defined as unchanged insulin requirement or glycemic control in 26% (n=19). Additionally, six of these 72 patients (8%) remained insulin-independent from diabetes onset until their median follow-up time of 1 year (3 days – 3 years) after initiation of thiamine. Three patients required insulin-treatment again at puberty. Only 11 of all 95 patients had reached adulthood (aged >18y)-all of these were on insulin therapy.

Glycemic control both prior to and after the initiation of thiamine treatment was reported for 29 of 95 (31%) patients with a median pre-HbA1c of 8.7% (5.9 – 21.0%) and median post-HbA1c 6.7% (5.2–9.1%)^89,93–94,100–103,105,109,110–115,117^. Notably, the reported pre-HbA1c was in most cases measured at diabetes onset, therefore a decrease in HbA1c does not alone reflect thiamine effect on glycemic control, but could also reflect the effect of standard treatment of diabetes. Insulin dose at both timepoints was available for 16 of 89 (18%) insulin-treated patients, with the median dose of 0.68 (0.42 – 1.8) IU/kg/d prior to thiamine treatment and 0.4 (0.0–0.8) IU/kg/d while on thiamine treatment.

The initial and maximum median thiamine doses were 100 (25 - 200) mg/d and 100 (25 – 600), respectively mg/d (n=41), adjusted according to the response on anemia.

## Discussion

This is the first systematic review to look at the evidence for precision treatment of beta-cell monogenic diabetes outside of K_ATP_ neonatal diabetes, in which SU were previously shown to be an effective treatment with the success conditioned by differences in pharmacogenetics, age, pharmacokinetics, compliance, and maximal dose used^5^. Monogenic diabetes is an excellent candidate for precision medicine as the genetic etiology identified by genetic testing defines a subtype of diabetes with a specific pathophysiology enhancing the likelihood for a specific therapy to be most effective. For several forms of beta-cell monogenic diabetes, the underlying pathoetiology provides a rationale for precision treatment, but it is important to assess to what extent these rationales are supported by published evidence.

We sought to assess the evidence base for current treatment recommendations for several more common forms of beta-cell monogenic diabetes. Overall we found that there is limited, and mostly poor quality evidence, mainly consisting of case reports and case series. There were limited randomized studies providing stronger evidence, some with considerable effect sizes, but limited by small numbers of participants and short durations. The strongest evidence for a precision approach was, in order, HNF1A-diabetes, GCK-related hyperglycemia, relapse of 6q24-TND and SLC19A2-diabetes. The evidence for insulin or non-insulin therapies in MD and HNF1B-diabetes was not clear and there was almost no evidence for HNF4A-diabetes treatment. Each subtype is discussed below.

### GCK-related hyperglycemia

The aggregate of data provide evidence that glucose lowering treatment should not be given in GCK-related hyperglycemia. HbA1c stays stable in target range without initiating or after cessation of medical treatment and there is no evidence to support treatment for lowering glycemia. However, diagnostic testing for GCK is often guided by recruiting individuals with mild hyperglycemia and this may introduce bias to the phenotype seen. Against this, in non-selected sequencing in the general population^120^ and type 2 diabetes^121^, a mild glycemic phenotype was seen in subjects with *GCK* variants.

There was no evidence to support dietary recommendations specific to GCK-related hyperglycemia. The one cross-over study of diet was not a trial of dietary advice, studied the glucose response to 2 days exposure to high and low carbohydrate and had methodological limitations^13^. Further work is needed on whether a dietary approach needs to be any different from the general population.

A limitation of this systematic review is that we did not address the evidence for a precision medicine approach in pregnancies affected by maternal beta-cell monogenic diabetes. This is an important area particularly in GCK-related hyperglycemia, where treatment may be required for pregnancies in women with GCK-related hyperglycemia carrying non-affected fetuses^122^. Management guidelines exist but are debated. Additionally, the potential impact of cell-free fetal DNA to provide early genotype information to guide maternal insulin therapy will need to be studied both in terms of treatment impact and cost-effectiveness. These are important topics for assessment in GCK-related hyperglycemia in the future.

*Recommendation: No medical treatment should be given in GCK-related hyperglycemia (grade C evidence). No recommendation can be given regarding dietary treatment in GCK-related hyperglycemia*.

### HNF1A-diabetes and HNF4A-diabetes

Notably almost all data available was for HNF1A-diabetes. Therefore, we can only provide assessment of the evidence in HNF1A-diabetes although combined cohorts suggest results may be similar in HNF4A-diabetes. Clearly more treatment studies are required in HNF4A-diabetes

The strongest evidence in this systematic review for precision therapy was for the use of SU in HNF1A-diabetes. The key evidence was a randomized cross-over study of SU and metformin therapy in HNF1A-diabetes and matched subjects with type 2 diabetes^24^. Treatment response to SU in HNF1A-diabetes was significantly greater than that seen in type 2 diabetes, establishing treatment response difference by genetic subtype. The aggregate of data supports the initial efficacy of SU at diabetes diagnosis and the possibility of transitioning from insulin therapy after a genetic diagnosis is made, especially when close to diagnosis and when the HbA1c is well controlled^22^. Further studies are needed to study factors that influence the response to SU within HNF1A-diabetes.

The effectiveness of glinides in HNF1A-diabetes as an alternative to SU, particularly to address hypoglycemia, is also supported albeit with limited data^26,30,44^. While the newer diabetes medications have demonstrated effectiveness as monotherapy (GLP1RA)^26,29,42^ or augmentative therapy (DPP4i)^20,27,31,33,36,38^, evidence for these treatments is limited and studies are of short duration. Moreover, treatment response has not been compared between HNF1A-diabetes and HNF4A-diabetes and type 2 diabetes as was tested with SU.

There are several remaining questions in the area of precision treatment for HNF1A-diabetes and HNF4A-diabetes. While SU are a clear representation of diabetes precision medicine, they are non-efficacious in some individuals and their efficacy may not be durable in others, with diabetes duration and weight gain being two factors associated with reduced SU efficacy. The increased cardiovascular risk associated with HNF1A-diabetes provides another reason that carefully-designed, long-term comparison studies of glycemic and cardiovascular outcomes of these newer classes of diabetes medications, which offer cardiovascular benefit, are needed^123,124^.

*Recommendation: SU should be used as first-line therapy for HNF1A-diabetes (grade C evidence). Glinides can be used if frequent hypoglycemia is experienced with SU treatment (grade D evidence). GLP1RA are an option in treatment of HNF1A-diabetes (grade C evidence). DPP4i are an option in augmentative treatment of HNF1A-diabetes (grade C evidence). No recommendation can be given for HNF4-diabetes*.

### HNF1B-diabetes and Mitochondrial diabetes

The evidence for specific treatment in HNF1B-diabetes and MD is of low quality with no trials and very few studies. The degree of insulin deficiency among the individuals with HNF1B-diabetes and MD can range from mild hyperglycemia to absolute insulin deficiency. While insulin has been advocated as the choice of treatment for both HNF1B-diabetes and MD^4,125^, we found no systematic evidence favoring the use of insulin. There are no RCTs on HNF1B-diabetes or MD and even open treatment studies and cohort or case reports are few. Thus, the results of the systematic search remain descriptive reflecting the choices of the treating physicians. In the included case reports and case series, which were mainly cross-sectional, most patients diagnosed with HNF1B-diabetes or MD seem to have commenced insulin treatment at some stage. Besides insulin deficiency and secondary failure of other medications, this could be affected by a diagnostic selection bias or a progressive kidney disease precluding many modes of treatment. Similarly, there is no evidence for using or avoiding any of the other diabetes medications. We note that metformin, SU or glinides, DPP4i, SGLT2i and GLP1RA were used in the case reports and case series especially close to diagnosis. Further large systematic and controlled studies are required in both HNF1B-diabetes and MD.

Lactatemia and/or lactic acidosis is part of the syndrome caused by the m.3243A>G variant^4^. Thus, avoidance of metformin has been recommended in MD as it impairs mitochondrial function and may further increase the risk of lactic acidosis^4^. However, evidence against its clinical use was low and limited to three case reports with adverse effects, including only one patient with possible lactic acidosis^67^ and another with an increase of lactate levels possibly associated with metformin use^65^. MD needs additional preclinical and clinical studies to define risk of metformin and statin use to guide therapy approaches. Patients with m.3243A>G and associated neurological disease might be on coenzyme Q10 or other dietary supplements, like arginine, in an attempt to support their mitochondrial function but most studies have not examined the effects of the supplements on glucose control.

*Recommendation: There is insufficient data to recommend or refute the preferential use of insulin or any other medication in HNF1B-diabetes or mitochondrial diabetes. However, in case of patients not on insulin therapy, potential deterioration of insulin secretion should be evaluated if the glucose control deteriorates*.

### 6q24 TNMD

The evidence guiding the use of non-insulin therapies for 6q24-related diabetes is of low quality and more information is needed to make firm conclusions. In the initial neonatal phase of 6q24-TND, SU use was not always successful in improving diabetes outcome or allowing for cessation of insulin. The evidence for efficacy of non-insulin therapies was stronger for the relapse phase of diabetes later in life, where most cases appeared to benefit from a variety of non-insulin therapies, with most not requiring insulin. There is a pressing need to improve the evidence base for management of 6q24-related diabetes in both the neonatal and relapse phases, and for long-term outcome data.

*Recommendation: There is insufficient data to recommend or refute the preferential use of non-insulin therapies for the treatment of the neonatal or relapse phase of 6q24-diabetes. However, non-insulin therapy seems beneficial in patients where diabetes recurs, but we recommend close follow-up to ensure treatment intensification if patients do not reach or remain stable at their glycemic target (grade D evidence)*.

### SLC19A2 diabetes (TRMA)

The evidence for use of thiamine improving glycemic control in SLC19A2-diabetes is limited by the lack of larger studies, long-term follow up, and randomized clinical trials. However, there is a consistent trend across the cohort and case studies reporting thiamine administration resulting in a reduction in insulin dose, including discontinuation in some cases, and improvement in glycemia. Quantifying an improvement in endogenous insulin secretion in patients treated with insulin is difficult but probably the best assessment is the daily insulin dose per kg in patients who maintain a similar or improved level of glycemia. Further trials assessing this in the short and long term (especially beyond puberty) are required to firmly document the effect of thiamine on glycemic control.

However, in contrast to the other forms of monogenic diabetes described in this review, the precision treatment thiamine, should be started early for the severe anemia in SLC19A2-diabetes and continued following establishment of thiamine-responsiveness. This special situation and the rarity of the disease make clinical trials difficult to conduct.

*Recommendation: Whereas thiamine treatment is essential for all patients with anemia in SLC19A2-diabetes, the evidence supporting a specific and sustained effect of thiamine on glycemic control in SLC19A2-diabetes is weak. We recommend in SLC19A2-diabetes that thiamine is started as soon as a diagnosis is made. Additionally, we recommend close follow up of patients with SLC19A2-diabetes after the initiation of thiamine treatment to adjust the diabetes treatment if required (grade D evidence)*.

### Concluding remarks

Beta-cell monogenic diabetes has been considered the strongest example of a precision medicine approach to diabetes treatment. However, this systematic review demonstrates that there is limited trial evidence to support precision treatment practices and so many recommendations rely on case series and case reports. Importantly the strongest available evidence support the existing practice consensus guidelines^4^ regarding GCK-related hyperglycemia and HNF1A-diabetes.

Randomized trials with long-term follow-up offer the strongest evidence but the low numbers of cases of each individual subtype make these very difficult to perform. We urge the medical community to publish the follow-up of complete case series in all subtypes of beta-cell monogenic diabetes to establish the response to therapy, fill the identified gaps in precision treatment and to explore the precision treatment approaches to prevent complications. Areas where we identify further work being required include:

- There are no studies on drug-naïve newly-diagnosed patients with HNF1A- or HNF4A-diabetes comparing the different treatment options. This would be needed to conclusively suggest first-line (or second-line) treatment of hyperglycemia.
- Given the increased cardiovascular risk in HNF1A-diabetes, larger studies of long duration with glycemic and cardiovascular endpoints comparing SU with cardioprotective GLP1RA or SGLT2i are warranted. However, carefully designed studies are needed as it is not clear whether SGLT2i can be safely used in HNF1A-diabetes, due to the degree of insulin deficiency and already reduced expression of SGLT2 and glycosuria as a feature.
- There is no defined precision treatment approach or even clear first-line medication for MD or HNF1B-diabetes. There is a rationale for studying SGLT2i specifically in HNF1B-diabetes, as it might improve the renal outcome of the associated non-diabetes related kidney disease. However, the marked insulin-deficiency often seen in HNF1B-diabetes could markedly increase the risk of diabetic ketoacidosis.
- While GCK-related hyperglycemia occurring in isolation does not need pharmacologic treatment, it is important to develop clinical recommendations for diagnosing and treating type 2 diabetes co-occurring with GCK-related hyperglycemia.
- Future studies of precision treatment in beta-cell monogenic diabetes must account for the perspectives of people with monogenic diabetes, including acceptability of the different medications in terms of route, patient-facing costs, and potential adverse effects. Cost-effectiveness analyses of the newer diabetes medications also need to be carried out. Finally, there must be purposeful efforts toward equity in achieving optimal diabetes outcomes for individuals living with diabetes, with special attention to groups of people and countries underrepresented in studies of monogenic diabetes.

## Authors’ declaration of personal interests

JK has received lecture fees from NovoNordisk, international conference costs covered by Medtronic, AstraZeneca and NovoNordisk; MP has been the scientific advisor for the AMGLIDIA (glibenclamide suspension) development; TV has served on scientific advisory panels, been part of speaker’s bureaus, and served as a consultant to, and/or received research support from Amgen, AstraZeneca, Boehringer Ingelheim, Eli Lilly, Gilead, GSK, Mundipharma, MSD/Merck, Novo Nordisk, Sanofi, and Sun Pharmaceuticals.

RNN, KAP, JMEM, JS, JB, SAWG, ATH, TT report no conflicts of interest.

## Supporting information

Supplemental tables of search strategy

## Data Availability

All data produced in the present study are available upon reasonable request to the authors.

## Acknowledgements

We acknowledge the contribution of Dr. Rebecca Brown and Dr. Robert Semple in the working group of the Treatment of Monogenic Diabetes of the ADA/EASD Precision Diabetes Medicine Initiative. R.N.N is supported by the following grants: ADA 7-22-ICTSPM-17; R01DK104942; U54DK118612. KAP is supported by Wellcome Trust (219606/Z/19/Z); M.P. is supported by ANR 22-CE17-0025 Neurogli; S.A.W.G is supported by NIH NIDDK R01DK104942 and U54DK118612; T.T. is supported by Folkhalsan Research Foundation as well as The Academy of Finland (grants no. 336822, 312072 and 336826) and University of Helsinki for the Centre of Excellence of Complex Disease Genetics.

The ADA/EASD Precision Diabetes Medicine Initiative, within which this work was conducted, has received the following support: The Covidence license was funded by Lund University (Sweden) for which technical support was provided by Maria Björklund and Krister Aronsson (Faculty of Medicine Library, Lund University, Sweden). Administrative support was provided by Lund University (Malmö, Sweden), University of Chicago (IL, USA), and the American Diabetes Association (Washington D.C., USA). The Novo Nordisk Foundation (Hellerup, Denmark) provided grant support for in-person writing group meetings (PI: L Phillipson, University of Chicago, IL).

## Author contributions

*Review Design*: RNN, KAP, JK, JS, SAWG, TV, ATH, TT

*Systematic Review Implementation*: RNN, KAP, JK, JMEM, JS, SAWG, TV, TT

*Data extraction manuscripts:* RNN, KAP, JK, JMEM, SAWG

*Manuscript writing:* RNN, KAP, JK, JMEM, JS, JB, MP, TV, SAWG, ATH, TT

*Project Management:* TV, SAWG, ATH, TT

