## Supplemental tables of search strategy for "Systematic Review of Treatment of Beta-Cell Monogenic Diabetes"

**Supplemental Tables: Search Strategies**

**Supplemental Table 1:** Search terms and keywords used to identify relevant studies for GCK-related hyperglycemia, HNF1A-diabetes and HNF4A-diabetes

| **Diabetes phenotypes** | **Gene Names** | **Treatments** | **Outcomes** |
| --- | --- | --- | --- |
| Monogenic diabetes | Hepatocyte Nuclear Factor 1 Alpha | Metformin | HbA1c |
| MODY | HNF1 | Biguanide | Treatment response |
| Maturity onset diabetes of the young | Hepatocyte Nuclear Factor 1-Alpha | Sulphonylurea | Glycemia |
|  | Transcription Factor HNF-1 | Sulfonylurea | Treatment failure |
|  | Hepatic Nuclear Factor 1 Alpha | Gliclazide | Intolerance |
|  | HNF1alpha | Glipizide | Adverse effect |
|  | HNF-1-Alpha | Glibenclamide | Side effect |
|  | HNF1alpha | Glyburide | Diarrhea |
|  | HNF1a | Glimepiride | Lactic acidosis |
|  | MODY3 | Tolbutamide | Acute kidney injury |
|  | HNF1A | Chlorpropamide | Hypoglycemia |
|  | TCF-1 | Repaglinide | Weight gain |
|  | TCF1 | Nateglinide | Edema |
|  | MODY-3 | Thiazolidinedione | Ketosis |
|  | Hepatocyte Nuclear Factor 4 Alpha | PPARg Agonist | Ketoacidosis |
|  | HNF4 | Rosiglitazone | Pancreatitis |
|  | Hepatocyte Nuclear Factor 4-Alpha | Pioglitazone | Nausea |
|  | Transcription Factor HNF-4 | Troglitazone | Vomiting |
|  | Hepatic Nuclear Factor 4 Alpha | Alphaglucosidase inhibitor | Cardiovascular disease |
|  | HNF4alpha | Acarbose | Myocardial infarction |
|  | HNF-4-Alpha | DPP4 inhibitor | Revascularisation |
|  | HNF4alpha | DPP-4 inhibitor | Acute coronary syndrome |
|  | HNF4a | Dipeptidylpeptidase 4 inhibitor | Coronary heart disease |
|  | MODY1 | Dipeptidylpeptidase-4 inhibitor | Heart failure |
|  | HNF4A | Sitagliptin | Cardiac Failure |
|  | TCF-14 | Vildagliptin | Angioplasty |
|  | TCF14 | Saxagliptin | Percutaneous Coronary Intervention |
|  | MODY-1 | Linagliptin | Cardiovascular Mortality |
|  | Glucokinase | Alogliptin | MACE |
|  | HK4 | SGLT2 inhibitor | Stroke |
|  | Glucokinase (Hexokinase 4) | SGLT2i | TIA |
|  | Hexokinase Type IV | SGLT-2 inhibitor | Nephropathy |
|  | Hexokinase-4 | SGLT-2i | Proteinuria |
|  | HK IV | Sodium Glucose Transporter 2 inhibitor | Macroalbuminuria |
|  | Hexokinase D, Pancreatic Isozyme | Dapagliflozin | Renal impairment |
|  | Hexokinase 4 | Empagliflozin | Chronic kidney disease |
|  | Hexokinase-D | Ertagliflozin | Microalbuminuria |
|  | MODY2 | Canagliflozin | Retinopathy |
|  | GCK | GLP1RA | Neuropathy |
|  | MODY-2 | GLP1 Receptor Agonists |  |
|  |  | GLP-1 Receptor Agonists |  |
|  |  | GLP-1RA |  |
|  |  | Exenatide |  |
|  |  | Liraglutide |  |
|  |  | Lixisenatide |  |
|  |  | Semaglutide |  |
|  |  | Dulaglutide |  |
|  |  | Albiglutide |  |
|  |  | Isophane |  |
|  |  | NPH insulin |  |
|  |  | Basal Insulin |  |
|  |  | Long acting insulin |  |
|  |  | Glargine |  |
|  |  | Detemir |  |
|  |  | Degludec |  |
|  |  | Insulin |  |
|  |  | Lifestyle |  |
|  |  | Diet |  |
|  |  | Dietary |  |
|  |  | Weight Loss |  |
|  |  | Exercise |  |
|  |  | Bariatric surgery |  |
|  |  | Obesity Surgery |  |
|  |  | Weight reduction surgery |  |
|  |  | gastric band |  |
|  |  | Roux-en-Y |  |
|  |  | Gastric sleeve |  |
|  |  | Gastric bypass |  |

Overall Search: (diabetes phenotype AND each gene AND treatment AND outcome) using US and UK spellings, from 1992 and English language only

**Supplemental Table 2:** Search terms and keywords used to identify relevant studies for HNF1B-diabetes

| **Diabetes Phenotypes** | **Genes** | **Treatments** |
| --- | --- | --- |
| MODY | HNF1 Homeobox B | Metformin |
| monogenic diabetes | vHNF1 | Biguanide |
| MODY5 | Hepatocyte Nuclear Factor 1-Beta | Sulphonylurea |
| MODY-5 | Hepatocyte Nuclear Factor 1β | Sulfonylurea |
| HNF1B-MODY | Hepatocyte Nuclear Factor 1-β | Gliclazide |
| HNF1B-diabetes | HNF-1-Beta | Glipizide |
| HNF1B diabetes | HNF1-β | Glibenclamide |
| HNF1B disease | HNF-1β | Glyburide |
| HNF1B-disease | HNF1beta | Glimepiride |
| HNF1B-related disease | HNF-1B | Tolbutamide |
| renal cysts and diabetes | MODY5 | Chlorpropamide |
| RCAD | TCF-2 | Repaglinide |
|  | TCF2 | Nateglinide |
|  | HNF1 Beta | Thiazolidinedione |
|  | HNF1B | PPARg Agonist |
|  | MODY-5 | Rosiglitazone |
|  | hepatocyte nuclear factor 1b | Pioglitazone |
|  | hepatocyte nuclear factor 1 beta | Troglitazone |
|  | hepatocyte nuclear factor 1 β | Alphaglucosidase inhibitor |
|  |  | Acarbose |
|  |  | DPP4 inhibitor |
|  |  | DPP-4 inhibitor |
|  |  | Dipeptidylpeptidase 4 inhibitor |
|  |  | Dipeptidylpeptidase-4 inhibitor |
|  |  | Sitagliptin |
|  |  | Vildagliptin |
|  |  | Saxagliptin |
|  |  | Linagliptin |
|  |  | Alogliptin |
|  |  | SGLT2 inhibitor |
|  |  | SGLT2i |
|  |  | SGLT-2i |
|  |  | Sodium Glucose Transporter 2 inhibitor |
|  |  | Dapagliflozin |
|  |  | Empagliflozin |
|  |  | Ertagliflozin |
|  |  | Canagliflozin |
|  |  | GLP1RA |
|  |  | GLP1 Receptor Agonists |
|  |  | GLP-1 Receptor Agonists |
|  |  | GLP-1RA |
|  |  | Exenatide |
|  |  | Liraglutide |
|  |  | Lixisenatide |
|  |  | Semaglutide |
|  |  | Dulaglutide |
|  |  | Albiglutide |
|  |  | Isophane |
|  |  | NPH insulin |
|  |  | Basal Insulin |
|  |  | Long acting insulin |
|  |  | Glargine |
|  |  | Detemir |
|  |  | Degludec |
|  |  | Insulin |
|  |  | Lifestyle |
|  |  | Diet |
|  |  | Dietary |
|  |  | Weight Loss |
|  |  | Exercise |
|  |  | Bariatric surgery |
|  |  | Obesity Surgery |
|  |  | Weight reduction surgery |
|  |  | gastric band |
|  |  | Roux-en-Y |
|  |  | Gastric sleeve |
|  |  | Gastric bypass |

Overall Search: (diabetes phenotype AND each gene AND treatment) using US and UK spellings, from 1992 and English language only

**Supplemental Table 3:** Search terms and keywords used to identify relevant studies for mitochondrial diabetes

| **Diabetes Phenotypes** | **Genes** | **Diabetes Treatments** | **Other Treatments 1** | **Other Treatments 2** |
| --- | --- | --- | --- | --- |
| MIDD* | MTTL1 | Metformin | statin(s) | Vitamin(s) |
| MELAS and (diabetes | Mitochondrially Encoded TRNA-Leu | Biguanide | fibrate(s) | dietary supplements |
| MELAS AND diabetes OR hyperglycemia OR hyperglycaemia OR dysglycemia OR dysglycaemia | Mitochondrially Encoded TRNA Leucine 1 | Sulphonylurea | PCSK9-inhibitor(s) | ubiquinol |
| mitochondrial diabetes | MT-TL1 | Sulfonylurea | ezetimibe | ubiquinone |
| maternally inherited diabetes | MTTE | Gliclazide | simvastatin | ubidecarenone |
| maternally inherited diabetes and deafness | MT-TE | Glipizide | rosuvastatin | coenzyme Q |
| maternal diabetes and deafness | Mitochondrially Encoded TRNA-Glu | Glibenclamide | atorvastatin | CoQ_10_ |
| Mitochondrial encephalopathy, lactic acidosis, and stroke-like episodes AND diabetes OR hyperglycemia OR hyperglycaemia OR dysglycemia OR dysglycaemia | Mitochondrially Encoded TRNA Glutamic Acid | Glyburide | pravastatin |  |
| Mitochondrial encephalopathy, lactic acidosis, and stroke like episodes AND diabetes OR hyperglycemia OR hyperglycaemia OR dysglycemia OR dysglycaemia | MTTK | Glimepiride | Fluvastatin |  |
|  | Mitochondrially Encoded TRNA-Lys | Tolbutamide |  |  |
|  | Mitochondrially Encoded TRNA Lysine | Chlorpropamide |  |  |
|  | MT-TK | Repaglinide |  |  |
|  | m.3243A | Nateglinide |  |  |
|  | mt.3243 | Thiazolidinedione |  |  |
|  | A3243G | PPARg Agonist |  |  |
|  | m.3243A>G | Rosiglitazone |  |  |
|  | mt.3243A>G | Pioglitazone |  |  |
|  | 3243A-G | Troglitazone |  |  |
|  | 3243 a to g | Alphaglucosidase inhibitor |  |  |
|  | m. 14709 | Acarbose |  |  |
|  | mt. 14709 | DPP4 inhibitor |  |  |
|  | T14709C | DPP-4 inhibitor |  |  |
|  | m. 14709T>C | Dipeptidylpeptidase 4 inhibitor |  |  |
|  | mt. 14709T | Dipeptidylpeptidase-4 inhibitor |  |  |
|  | 14709T-C | Sitagliptin |  |  |
|  | 14709 t to c | Vildagliptin |  |  |
|  | m.8396 | Saxagliptin |  |  |
|  | mt.8396 | Linagliptin |  |  |
|  | A8396G | Alogliptin |  |  |
|  | m.8396A | SGLT2 inhibitor |  |  |
|  | mt.8396A>G | SGLT2i |  |  |
|  | 8396A-G | SGLT-2i |  |  |
|  | 8396 a to g | Sodium Glucose Transporter 2 inhibitor |  |  |
|  | 3243 | Dapagliflozin |  |  |
|  | 14709 | Empagliflozin |  |  |
|  | 8396 | Ertagliflozin |  |  |
|  |  | Canagliflozin |  |  |
|  |  | GLP1RA |  |  |
|  |  | GLP1 Receptor Agonists |  |  |
|  |  | GLP-1 Receptor Agonists |  |  |
|  |  | GLP-1RA |  |  |
|  |  | Exenatide |  |  |
|  |  | Liraglutide |  |  |
|  |  | Lixisenatide |  |  |
|  |  | Semaglutide |  |  |
|  |  | Dulaglutide |  |  |
|  |  | Albiglutide |  |  |
|  |  | Isophane |  |  |
|  |  | NPH insulin |  |  |
|  |  | Basal Insulin |  |  |
|  |  | Long acting insulin |  |  |
|  |  | Glargine |  |  |
|  |  | Detemir |  |  |
|  |  | Degludec |  |  |
|  |  | Insulin |  |  |
|  |  | Lifestyle |  |  |
|  |  | Diet |  |  |
|  |  | Dietary |  |  |
|  |  | Weight Loss |  |  |
|  |  | Exercise |  |  |
|  |  | Bariatric surgery |  |  |
|  |  | Obesity Surgery |  |  |
|  |  | Weight reduction surgery |  |  |
|  |  | gastric band |  |  |
|  |  | Roux-en-Y |  |  |
|  |  | Gastric sleeve |  |  |
|  |  | Gastric bypass |  |  |

*Not monoclonal Ig deposition disease, model informed drug development

Overall Search: (diabetes phenotype and genetic variant AND (diabetes treatment OR Other Treatments 1 OR Other Treatments 2) using US and UK spellings, from 1992 and English language only

**Supplemental Table 4:** Search terms and keywords used to identify relevant studies for 6q24 transient neonatal diabetes

| **Diabetes Phenotypes** | **Genes** |
| --- | --- |
| Diabetes Mellitus, Permanent Neonatal | PLAGL1 protein, human |
| Diabetes Mellitus, Transient Neonatal, 3 | HYMAI, RNA |
| 6q24-Related Transient Neonatal Diabetes Mellitus | PLAGL1 |
| Diabetes Mellitus, Neonatal, with Congenital Hypothyroidism | HYMAI |
| Diabetes Mellitus, Permanent Neonatal, with Cerebellar Agenesis | ZPF57 |
| Diabetes Mellitus, Transient Neonatal, 2 | 6q24 |
| Diabetes Mellitus, Transient Neonatal, 1 | chromosome 6q24 |
| maturity-onset diabetes of the young | UPD6 |
| MODY | Uniparental Disomy of Chromosome 6 |
| diabetes mellitus/genetics |  |
| monogenic diabetes |  |
| neonatal diabetes |  |
| KCNJ11-diabetes |  |
| ABCC8-diabetes |  |
| KCNJ11-PNDM |  |
| ABCC8-PNDM |  |
| infancy-onset diabetes |  |
| NDM |  |
| PNDM |  |
| TNDM |  |
| 6q24-related diabetes mellitus |  |
| 6q24-related diabetes |  |
| maturity onset diabetes |  |
| maturity-onset diabetes |  |
| MODY |  |
| 6q24-TNDM |  |
| 6q24 TNDM |  |
| iTND |  |
| DMTN |  |

Overall Search: (diabetes phenotype AND each gene) between 01/01/1999 – 06/01/2022 in humans and English language only

**Supplemental Table 5:** Search terms and keywords used to identify relevant studies for SLC19A2-diabetes

| **Diabetes Phenotypes** | **Genes** |
| --- | --- |
| Thiamine responsive megaloblastic anemia syndrome | SLC19A2 |
| megaloblastic anaemia and deafness | solute carrier family 19 (thiamine transporter), Member 2 |
| thiamine-responsive anemia | thiamine transporter 1 |
| Thiamine-responsive megaloblastic anemia | ThTr-1 |
| thiamine-responsive anemia | THTR1 |
| Rogers syndrome | THT1 |
| TRMA | THT-1 |
| Thiamine-responsive megaloblastic anaemia syndrome | TRMA |
| diabetes mellitus and sensorineural deafness and megaloblastic anemia | TC1 |
| diabetes mellitus and sensorineural deafness | TC-1 |
| megaloblastic anaemia | reduced folate carrier protein (RFC) like |
| Thiamine-responsive megaloblastic anemia syndrome with the addition | high affinity thiamine transporter |
| thiamine-responsiv* | thiamine carrier 1 |
| thiamine responsiv* | THMD1 |
| thiamineresponsiv* | THMD-1 |
| roger's syndrome* | ThTr1 |
| roger's disease* | THTR-1 |
| thiamine-dependent | thiamine transport |
| thiaminedependent | solute carrier family 19 (thiamine transporter), Member 2 |
| rogers syndrome* | SLC19A2 protein, human |
| rogers disease* |  |
| abboud disease* |  |
| abboud syndrome* |  |
| thiamine transport and metabolism |  |

Overall Search: (diabetes phenotype AND each gene) between 01/01/1999 – 06/01/2022 in humans and English language only
